# Assessing the impact of SARS-CoV-2 lineages and mutations on patient survival

**DOI:** 10.1101/2022.07.07.22277353

**Authors:** Carlos Loucera, Javier Perez-Florido, Carlos S. Casimiro-Soriguer, Francisco M. Ortuño, Rosario Carmona, Gerrit Bostelmann, L. Javier Martínez-González, Dolores Muñoyerro-Muñiz, Román Villegas, Jesus Rodriguez-Baño, Manuel Romero-Gomez, Nicola Lorusso, Javier Garcia-León, Jose M. Navarro-Marí, Pedro Camacho-Martinez, Laura Merino-Diaz, Adolfo de Salazar, Laura Viñuela, The Andalusian COVID-19 sequencing initiative, Jose A Lepe, Federico Garcia, Joaquin Dopazo

## Abstract

After more than two years of COVID-19 pandemic, SARS-CoV-2 still remains a global public health problem. Successive waves of infection have produced new SARS-CoV-2 variants with new mutations whose impact on COVID-19 severity and patient survival is uncertain. A total of 764 SARS-CoV-2 genomes sequenced from COVID-19 patients, hospitalized from 19th February 2020 to 30st April 2021, along with their clinical data, were used for survival analysis. A significant association of B.1.1.7, the alpha lineage, with patient mortality (Log Hazard ratio LHR=0.51, C.I.=[0.14,0.88]) was found upon adjustment by all the covariates known to affect COVID-19 prognosis. Moreover, survival analysis of mutations in the SARS-CoV-2 genome rendered 27 of them significantly associated with higher mortality of patients. Most of these mutations were located in the S, ORF8 and N proteins. This study illustrates how a combination of genomic and clinical data provide solid evidence on the impact of viral lineage on patient survival.

## Introduction

With more than 10 million sequences submitted to GISAID [1] and other databases, SARS-CoV-2 is probably one of the most widely sequenced pathogens. Successive waves of infection have resulted in a constant selection of SARS-CoV-2 variants with new mutations in their viral genomes [2-4]. Sometimes, these novel variants carry specific mutations that have been linked to higher transmissibility [5,6] and/or immune evasion [7,8], making them relevant from a public health perspective[9] and leading to their classification as variants of interest (VOI) or variants of concern (VOC)[10]. However, current studies have failed to provide solid evidences on the potential effects of viral variants or mutations on COVID-19 severity or patient survival. Paradoxically, the impact of host genetics over COVID-19 progression and patient survival [11], as recently revealed in case-controls [12] genome-wide association studies [13-15] and whole-genome sequencing studies [16], is better known than the impact of the viral variants or the mutations present in the viral genome. For example, while some studies suggest that lineages as B.1.1.7 (alpha) are associated with increased mortality [17] others could not find such association [18,19]. Epidemiological studies suggest that certain mutations, like the D614G mutation in the S protein, could be associated with higher mortality[20]. Studies using undetailed patient outcomes (with no covariates considered) find some mutations potentially associated with severe COVID-19[21]. Previously, a 382-nucleotide deletion in the open reading frame 8 was associated with a milder infection [22]. Actually, the definition or variants of concern (VOC) or variants of interest (VOI) is proposed by the World Health Organization (WHO)[10], the Centers for Disease Control and Prevention (CDC) [23] and COVID-19 Genomics UK Consortium (COG-UK) [24] are based on observed transmissibility, more severity in the disease or in vitro evidences of reduced antibody neutralization [25]. The phenotype of these VOCs and VOIs depend on the presence of specific mutations, known as mutations of concern[26], found to be associated with higher transmissibility [5,6] and/or immune evasion [7,8]. However, because of the lack of large datasets in which viral genomes and detailed patient clinical data are simultaneously available, studies providing solid evidence on the effects of viral variants or mutations on COVID-19 severity or patient survival are scarce. Thus, the use of large clinical data repositories in combination with systematic viral genome sequencing is an urgent need to determine these relationships of high relevance in clinics.

Andalusia, located in the south of Spain, is the third largest region in Europe, with a population of 8.4 million, equivalent to a medium-sized European country like Austria or Switzerland. In the beginning of the pandemics, Andalusia implemented an early pilot project for first-wave SARS-CoV-2 sequencing[27], which was later transformed into the Genomic surveillance circuit of Andalusia[28,29], a systematic genomic surveillance program in coordination with the Spanish Health Authority. In addition, the Andalusian Public Health System has systematically been storing the EHRs data of all Andalusian patients in the Population Health Base (BPS, acronym from its Spanish name “base poblacional de salud”) [30] since 2001, making of this database one of the largest repositories of highly detailed clinical data in the world (containing over 13 million of comprehensive registries) [30]. Data generated in both sequencing initiatives along with clinical data stored in BPS were used to carry out this study.

## Materials and Methods

### Design and patient selection

Among the whole genome SARS-CoV-2 sequences obtained from both, the pilot project of SARS-CoV-2 sequencing [27] (in which 1000 viral genomes corresponding to the first wave, randomly sampled, representative of all the COVID-19 diagnosis in Andalusia between 19^th^ February and 30^st^ June, 2020, were sequenced), the Spanish Genomic epidemiology of SARS-CoV-2 (SeqCOVID) [31] and the Genomic surveillance circuit of Andalusia[28,29] (that includes 2438 SARS-CoV-2 genomes corresponding to the second wave, systematic sequenced among PCR positive individuals, following the recommendations of the Spanish Ministry of Health [32]), a total of 764 of them corresponded to individuals which were hospitalized between 19^th^ February 2020 and 30^st^ April 2021. In particular, 287 samples corresponded to the pilot project, 103 to the SeqCIVID project and 374 to the sequencing circuit.

### Sequencing SARS-CoV-2 genome

SARS-CoV-2 RNA-positive samples were subjected to whole-genome sequencing at the sequencing facilities of the Genyo (Granada, Spain), Hospital San Cecilio (Granada, Spain), Hospital Virgen del Rocío (Sevilla Spain), IBIS (Sevilla, Spain) and CABIMER (Sevilla, Spain).

RNA preparation and amplification were carried out as described in the protocols published by the ARTIC network [33] using the V3 version of the ARTIC primer set from Integrated DNA Technologies (Coralville, IA, USA) to create correlative amplicons covering the SARS-CoV-2 genome, after cDNA synthesis using SuperScript IV Reverse Transcriptase (Thermo Fisher Scientific, Waltham, MA USA) 1 µl of random hexamer primers and 11 µL of RNA. Libraries were prepared according to the COVID-19 ARTIC protocol v3 and Illumina DNA Prep Kit (Illumina, San Diego, CA). Library quality was confirmed using Bioanalyzer 2100 system (Agilent Technologies, Santa Clara, CA). The libraries were then quantified by Qubit DNA BR (ThermoFisher Scientific, Waltham, MA, USA), normalized, and pooled, and sequencing was performed using Illumina MiSeq v2 (300 cycles) and NextSeq 500/550 Mid Output v2.5 (300 Cycles) sequencing reagent kits.

### Sequencing data processing

Sequencing data were analyzed using in-house scripts and the nf-core/viralrecon pipeline software [34]. Briefly, after read quality filtering, sequences for each sample are aligned to the SARS-CoV-2 isolate Wuhan-Hu-1 (GenBank accession: MN908947.3) [35] using bowtie2 algorithm [36], followed by primer sequence removal and duplicate read marking using iVar [37] and Picard [38] tools, respectively. Genomic variants were identified through iVar software, using a minimum allele frequency threshold of 0.25 for calling variants and a filtering step to keep variants with a minimum allele frequency threshold of 0.75. Using the set of high confidence variants and the MN908947.3 genome, a consensus genome per sample was finally built using iVar.

With the aim of having all the genomic variants of our dataset the whole set of consensus genomes, regardless of missing data, have been used as input to the Nextclade software [39]. Consensus genome was aligned against the SARS-CoV-2 reference genome and aligned nucleotide sequences were compared with the reference nucleotide sequence, one nucleotide at a time. Mismatches between the query and reference sequences are reported differently, depending on their nature: nucleotide substitutions, nucleotide deletions or nucleotide insertions. Lineage assignment to each consensus genome was generated by Pangolin tool [40].

The evolutionary rate of the virus was obtained using the Augur application [41]. Augur functionality relies on the IQ-Tree software [42], which estimates the phylogenetic tree by maximum likelihood using a general time reversible model with unequal rates and unequal base frequencies [43], from which the evolutionary rate is inferred.

### Clinical Data preprocessing

The Ethics Committee for the Coordination of Biomedical Research in Andalusia approved the study “Retrospective analysis of all COVID-19 patients in the entire Andalusian community and generation of a prognostic predictor that can be applied preventively in possible future outbreaks” (29th September, 2020, Acta 09/20). Clinical data for 764 hospitalized patients was requested from the BPS. The data were transferred from BPS to the Infrastructure for secure real-world data analysis (iRWD) [44] from the Foundation Progress and Health, Andalusian Public Health System.

The main primary outcome was COVID-19 death (certified death events during hospitalization). Following previous similar studies, the first 30 days of hospital stay were considered for survival calculations [45]. The time variable in the models corresponds to the length (in days) of hospital stay. Stays that imply one or more changes of hospital units are combined in a single stay where the admission and discharge dates are set to the start of the first and the end of the last combined stay, respectively. Finally, in order to reduce possible confounding effects due to reinfection mechanisms we have opted to include only the first stay for each patient. The data used from BPS to properly account for covariates known to be related with COVID-19 survival is listed in Table 1.

**Table 1.** Data imported from BPS for each patient: code and definition of the variable.

| **Code** | **Meaning** |
| --- | --- |
| FECNAC | Birth date |
| FECDEF | Death date |
| SEXO | Gender |
| FEC_INGRESO | Hospital admission date |
| FEC_ALTA | Discharge date |
| MOTIVO_ALTA | Reason for the discharge:<br>(recovery/death/admission in another<br>hospital/voluntary discharge/retirement<br>home/unspecified) |
| COD_PATOLOGIA_CRONICA | Hospital codes for chronic conditions |
| COD_FEC_INI_PATOLOGIA | Date of condition diagnosis |
| COD_CIE_NORMALIZADO | A mixture of ICD9 and ICD10 codes for diseases |
| DESC_CIE_NORMALIZADO | Description of the ICD |
| FECINI_DIAG | Diagnosis date |
| FECFIN_DIAG | End of the diagnosed condition |
| FUENTE_DIAG | Source of the diagnosis (hospital, emergency,<br>etc.) |
| IND_CRONICO_HCUP | Is it a chronic disease? (yes/no) |
| Test COVID: FECHA | Test COVID date |
| Test COVID: TYPE | PCR / antigens |
| Test COVID: RESULTADO_TEST | Result of the test (positive/negative)<sup>1</sup> |
| Pharmacy (Hospital and<br>external): DESCRIPCION | List of drugs used in hospital or purchased in the<br>pharmacies |
| Pharmacy (Hospital and<br>external): FECHA | Dispensing date |
| VACUNA | List of vaccines |
| VACUNAFECHA | Vaccination dates |

### Statistical analysis

The statistical analysis has been carried on both, at the level of lineages and at the level of mutations in the viral genome. In order to elucidate the association between each lineage / mutation and the survival outcome the following steps have been used: i) as a first step a covariate balance analysis to determine the viability for a causal analysis was applied [46]; ii) for these lineages or mutations suitable for causal analysis hazard ratios were obtained using the closed form variance estimator for weighted propensity score estimators with survival outcome [47]; iii) Also, a causal bootstrapped hazard ratio is obtained for the same lineages or mutations [48].

In detail, the first step involved the use of inverse probability weighting (IPW) for each mutation/lineage. IPW is based on propensity scores generated using the *WeightIt* R package (v 0.12) [49], where the exposed condition is, in the case of lineages, being infected by a virus of a specific lineage and, in the case of viral mutations, being infected by a virus harboring a specific mutation. To assess the viability of a causal analysis based on IPW the proportion of covariates that could be effectively balanced using the standardized mean differences test as implemented in the *Cobalt* R package (v 4.3.1) [50], was checked using the 0.05 threshold [46]. As covariates, variables previously associated with COVID-19 mortality were used, such as: age, sex, pneumonia/flu vaccination status, chronic obstructive pulmonary disease, hypertension, obesity, diabetes, chronic pulmonary and digestive diseases, asthma, chronic heart diseases and cancer [51] (see Table 1).

Covariate-adjusted Log hazard ratios (LHR) are computed for each mutation/lineage of interest using the closed form estimator as implemented in the *hrIPW* R package (v 0.1.3) [47]. For each analysis an estimate of the LHR along with a 95% confidence interval and a p value of significance was provided. This methodology provides a robust estimation of the variability of the LHR under IPW-bases tests [47].

A mutation or a lineage is considered eligible for a causal analysis if the closed form estimator converges and all the covariates could be properly balanced.

In addition, a bootstrapped estimation of the covariate-adjusted LHR has been computed, where the causal adjustment has been done using IPW as follows: i) the weights are computed with a binomial linear model where the response is the presence/absence of a given variant and the regressors are the covariates using *WeightIt*. ii) a Cox Proportional Hazards model as implemented in the R package *Survival* (v 3.2, iii) a bootstrapped 95% confidence interval of the LHR coefficient was computed using adjusted bootstrap percentile (BCa) as implemented in the R *Boot* package (v 1.3).

The theoretical p-values [47] associated with the survival outcome have been adjusted using the FDR method [52].

### Visualization of lineage prevalence over time

A script based in the CoVariants application [53] was used to visualize the distribution of lineage relative prevalence over the time period studied. Data from neighboring European countries (France, UK and Portugal) and Spain were taken from GISAID [54].

## Results

Here we used viral genomes from the pilot project of SARS-CoV-2 sequencing [27], the Genomic surveillance circuit of Andalusia [29] and the Spanish SeqCOVID project [31]. Among the individuals for which a SARS-CoV-2 whole genome sequence was available, 764 had a hospitalization event during the studied period, covering 19th February 2020 to 31st April 2021. According to PANGO lineage classification [55] a total of 18 SARS-CoV-2 lineages were identified among the 764 viral sequences used in this study (see Supplementary Table S1), 5 of them were eligible for causal analysis (see Methods): A, A.2, B.1, B.1.177 and B.1.1.7 (see Table 2). Figure 1 shows the circulation of different lineages in Andalusia and Spain during the period studied, and Supplementary Figure S1 shows the circulation in neighboring European countries. Although the different lineages emerged and declined approximately at the same time, documenting a fast and effective inter-country transmission, there are quantitative differences in their proportions. For example, B1.1.177 was far more prevalent in Spain and Andalusia that in the surrounding countries (Portugal, France and UK, see Supplementary Figure S1). However, the fast substitution of the alpha lineage (B.1.1.7) was similar in all the countries.

**Table 2:** pangolin lineages eligible for causal analysis and their equivalence with other nomenclatures.

| Pangolin lineage | Nextclade clade | WHO label |
| --- | --- | --- |
| A | 19B | - |
| A.2 | 19B | - |
| B.1 | 20A | - |
| B.1.177 | 20E | - |
| B.1.1.7 | 20I | Alpha |

**Figure 1.**
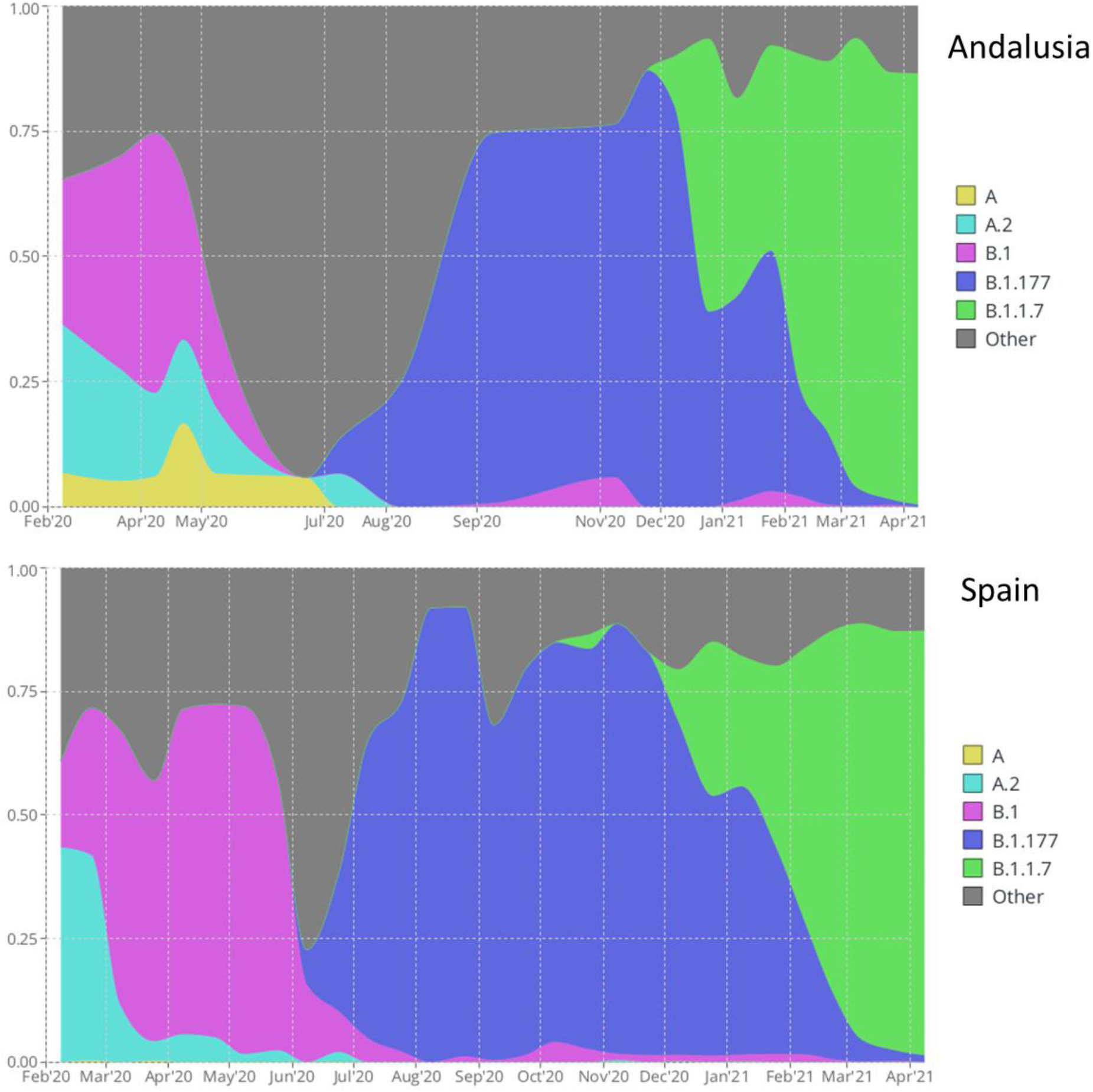
Circulation of the five SARS-CoV-2 variants eligible for the causal analysis in Andalusia (upper panel) and Spain (lower panel)

Figure 2 shows the log hazard ratios obtained for the different lineages. Only one of them, the B.1.1.7, has rendered a significant impact on patient survival (log Hazard Ratio, LHR, of 0.51, with a confidence interval C.I. = [0.14,0.88]). Interestingly, the A lineage, now virtually extinct, seems to cause a lower mortality than other lineages, although the result does not arrive to be significant (LHR= -1.80, C.I. = [-3.84,0.19]). However, retrospective survival analysis of lineages reveals relevant information on many lineages already extinct, or with very low representation, which limits its practical clinical application.

**Figure 2.**
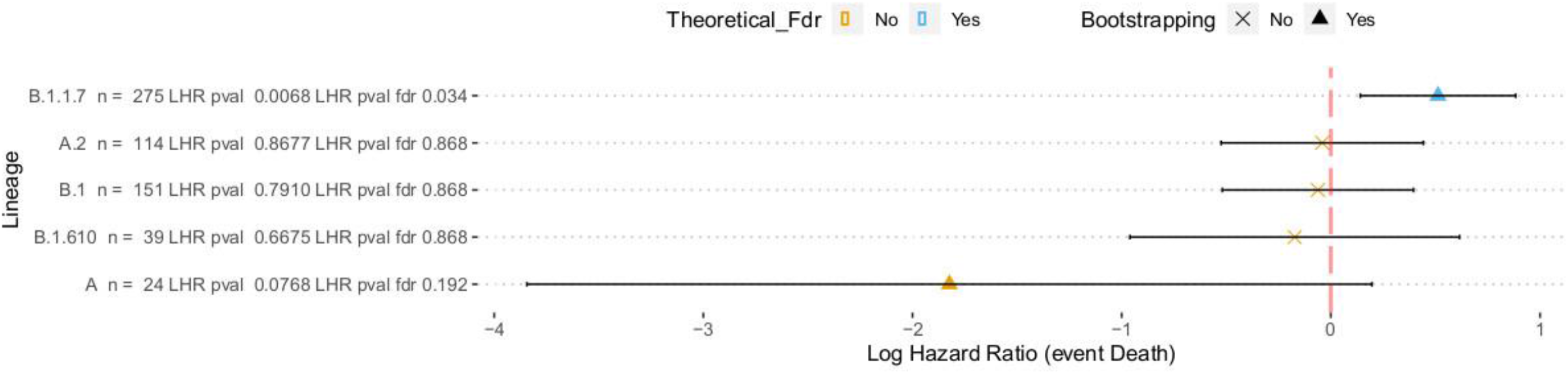
Log Hazard ratios estimated for the five variants eligible for the causal analysis using the two approaches described in the text (the closed form estimator and the bootstrap). For each analysis an estimate of the LHR along with a 95% confidence interval and a p-value (FDR adjusted) of significance is provided.

Contrarily, survival analysis of mutations provides interesting information, given that a large proportion of the studied mutations are still present in current lineages. Moreover, it throws light on regions of the proteins whose mutation could be related to higher mortality. A total of 594 nucleotide mutations were found with respect to the SARS-CoV-2 reference genome [35], 49 of which were eligible for formal causal analysis (see Methods). Figure 3 represents the LHR of the different mutations, plotted along the structure of the protein (see also Supplementary Figure S2). Among them, a total of 27 mutations presented a significant (FDR-corrected) association with patient survival, two of which have not been confirmed by subsequent bootstrapping analysis. Eighteen of them affect known Pfam [56] motifs (Table 3), some of them related to relevant viral features. For example, S:T716I affects the PF01601 motif (Coronavirus spike glycoprotein S2), which drives membrane penetration and virus-cell fusion and is involved in host specificity [57]; ORF8:Y73C, ORF8:R52I and ORF8:Q27*that affect the PF12093 (Betacoronavirus NS8 protein) motif, which allows SARS-CoV-2 ORF8 form unique large-scale assemblies which potentially mediate unique immune suppression and evasion activities [58,59]; or S:N501Y affects the PF09408 motif (Betacoronavirus spike glycoprotein S1, receptor binding), which has been implicated in binding to host receptors [60]. However, some motifs disrupted by mutations are of unknown function, as PF19211 or PF12379, corresponding to NSP2 and NSP3 proteins, respectively, which suggests that other relevant viral functionalities not yet characterized could be affected. Moreover, one of the significant mutations, ORF1ab:I2230T, does not affect to any known motif but it is significantly associated to patient higher mortality (see Figure 2 and Supplementary Table S2) by itself, given that it does not present correlations with other mutations (see Supplementary Figure S3). It is worth noting that some of these mutations associated to higher mortality in hospitalized unvaccinated patients are present in the current omicron variant, like ORF1ab:del3674-3676, S:del69-70 and S:del144 in BA.1 and S:N501Y and S:P681H in BA.1 and BA.2.

**Table 3.** Mutations associated with higher patient mortality that affect PFAM motifs and pangolin lineages eligible for caudal analysis with the mutation (non-synonymous from outbreak.info [63] and for synonymous from cov-spectrum.org [64].

| Mutation | position | cds | AAc position | AAc mutation | PFAM<sup>1</sup> | Definition | Lineages eligible for causal analysis bearing the mutation |
| --- | --- | --- | --- | --- | --- | --- | --- |
| C3267T | 3267 | ORF1ab | 1001 | ORF1ab:T1001I | PF12379 | Betacoronavirus replicase NSP3, N-terminal | A; B.1.177; B.1.1.7 |
| A4964G | 4964 | ORF1ab | 1567 | ORF1ab:T1567A | PF08715 | Coronavirus papain-like peptidase | B.1; B.1.1.7 |
| C5388A | 5388 | ORF1ab | 1708 | ORF1ab:A1708D | PF08715 | Coronavirus papain-like peptidase | B.1; B.1.177; B.1.1.7 |
| del11288.11297 | 11288 | ORF1ab | 3975-3677 | ORF1ab:del3675-3677 | PF08717 | Coronavirus replicase NSP8 | A; A.1; B.1; B.1.177; B.1.1.7 |
| C14676T | 14676 | ORF1ab | 4803 | ORF1ab:P4803P | PF00680 | RNA-dependent RNA polymerase | B.1; B.1.177; B.1.1.7 |
| C15279T | 15279 | ORF1ab | 5004 | ORF1ab:H5004H | PF00680 | RNA-dependent RNA polymerase | B.1; B.1.177; B.1.1.7 |
| del21766.21772 | 21766 | S | 69-70 | S:del69-70 | PF16451 | Betacoronavirus-like spike glycoprotein S1, N-terminal | A; A.1; B.1; B.1.177; B.1.1.7 |
| del21994.21997 | 21994 | S | 144 | S:Y144- | PF16451 | Betacoronavirus-like spike glycoprotein S1, N-terminal | A; A.1; B.1; B.1.177; B.1.1.7 |
| A23063T | 23063 | S | 501 | S:N501Y | PF09408 | Betacoronavirus spike glycoprotein S1, receptor binding | A; A.1; B.1; B.1.177; B.1.1.7 |
| C23271A | 23271 | S | 570 | S:A570D | PF19209 | Coronavirus spike glycoprotein S1, C-terminal | A; B.1; B.1.177; B.1.1.7 |
| C23709T | 23709 | S | 716 | S:T716I | PF01601 | Coronavirus spike glycoprotein S2 | B.1; B.1.177; B.1.1.7 |
| T24506G | 24506 | S | 982 | S:S982A | PF01601 | Coronavirus spike glycoprotein S2 | B.1; B.1.177; B.1.1.7 |
| G24914C | 24914 | S | 1118 | S:D1118H | PF01601 | Coronavirus spike glycoprotein S2 | B.1; B.1.177; B.1.1.7 |
| C27972T | 27972 | ORF8 | 27 | ORF8:Q27* | PF12093 | Betacoronavirus NS8 protein | A; B.1; B.1.177; B.1.1.7 |
| G28048T | 28048 | ORF8 | 52 | ORF8:R52I | PF12093 | Betacoronavirus NS8 protein | A; B.1; B.1.177; B.1.1.7 |
| A28111G | 28111 | ORF8 | 73 | ORF8:Y73C | PF12093 | Betacoronavirus NS8 protein | A; B.1; B.1.177; B.1.1.7 |
| C28977T | 28977 | N | 235 | N:S235F | PF00937 | Coronavirus nucleocapsid | A; B.1; B.1.177; B.1.1.7 |

<sup>1</sup> PFAM information can be accessed at: https://pfam.xfam.org/family/PFXXXX with PFXXXX being the corresponding PFAM ID

**Figure 3.**
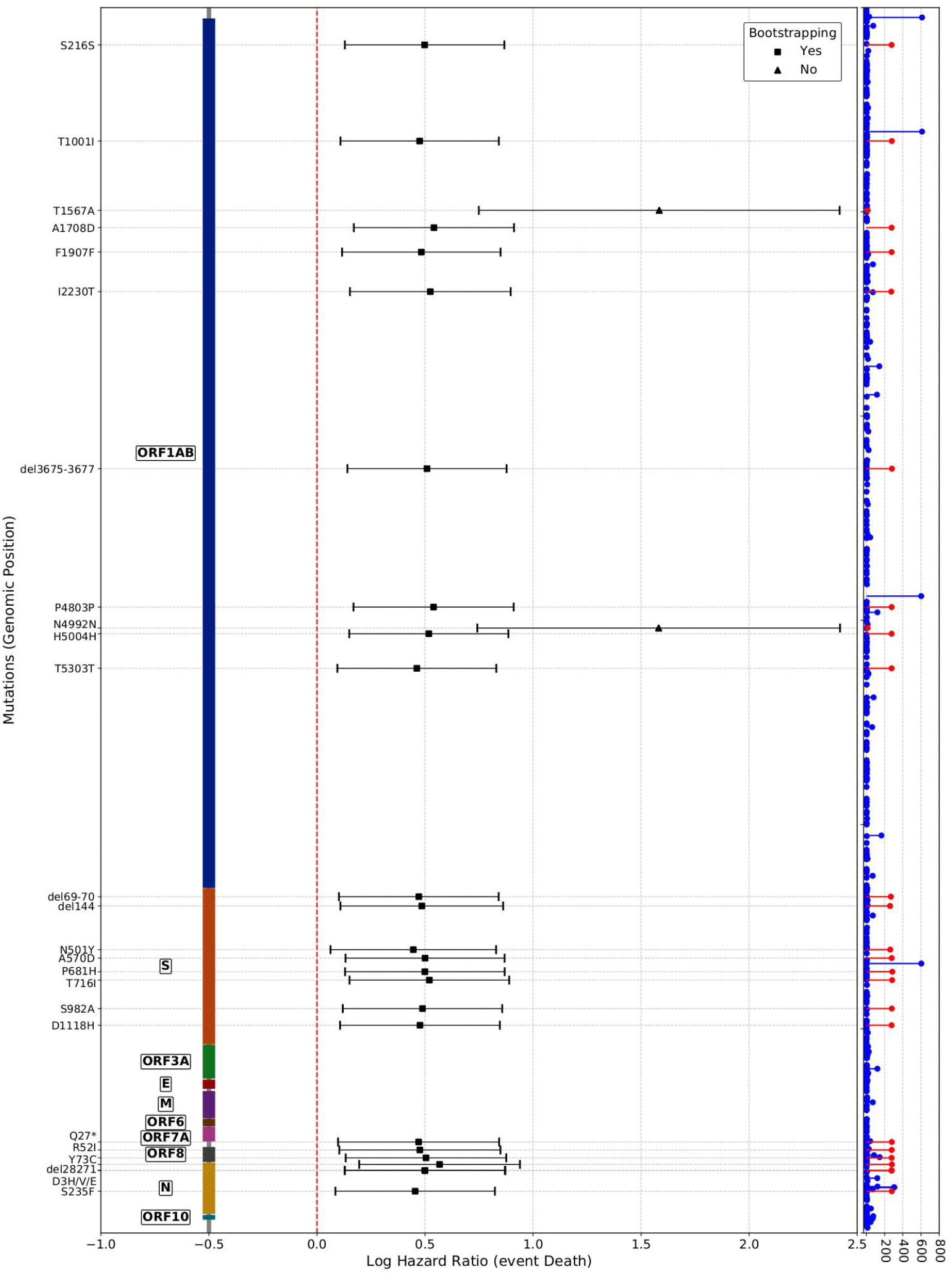
Log Hazard ratios estimated for the 25 viral mutations that presented a significant (FDR corrected) association with patient survival. Causal analysis was carried out using the two approaches described in the text (the closed form estimator and the bootstrap). For each analysis an estimate of the LHR along with a 95% confidence interval and a p-value (FDR adjusted) of significance is provided. Mutations are represented over the genomic positions in which they occur and on the left part, the corresponding proteins are annotated. The right part represents the observed distribution of mutations observed in all the samples analyzed.

Interestingly, some mutations in the viral genome seem to display a positive association with patient survival. The most remarkable case is the mutation ORF1ab:A3523V, which is significant with the bootstrap test (See Supplementary Table S2), although failed to be significant with the covariate-adjusted LHR test, because of the relatively small sample size.

The interest on mutations has focused mainly on non-synonymous changes, which produce a modification of the protein sequence that may have a potential influence on SARS-CoV-2 phenotypic properties. Contrarily, much less attention has been paid to synonymous changes, with a less clear relation with viral phenotypes, and currently there have been no reports of synonymous mutations of concern [25]. Here, for the first time, we describe nine synonymous mutations (G4300T, C2710T, C14676T, C15279T, C913T, C6968T, C5986T, C15240T and T16176C) in the ORF1ab with a significant association to higher mortality in COVID-19 hospitalized patients (Figure 3). However, some of them can simply be highly correlated with other coding mutations (e.g. C15279T is highly correlated with ORF1ab:T5303T, or C15240T is correlated with ORF1ab:T1567A) as depicted in Supplementary Figure S2.

The evolutionary rate displayed by the SARS-CoV-2 since February 2020 in the Andalusia region according to the SARS-CoV-2 whole genome sequencing circuit [28,29] is of 0.00063 substitutions per nucleotide per year (s/n/y), in agreement with the evolutionary rate previously described that ranges from 0.0004 and 0.002 s/n/y [2,4,25,61,62]. Interestingly, when mutations associated to high mortality (such as ORF1ab: A1708D) are depicted over the clock-adjusted phylogeny these tend to appear in the variants that have shaped the evolution of the virus in the Andalusia region during the period under study, many of them related to the alpha (B.1.1.7) lineage (See Supplementary Figure S4 A and B). The mutation associated with the highest mortality (ORF1ab:T1567A) shows a similar evolutionary rate (See Supplementary Figure S4 C) and it seems to define a specific clade within the alpha lineage (Supplementary Figure S4 D). However, some specific mutations, like the ones marginally associated to better survival (e.g. N:D377Y), appear in variants with apparently slower mutation rates (B.1.177, and sublineages), although it also appears in lineages B.1 and A,2, now extinct, and in a few variants, which are ancestors of the delta lineage. Actually, all the sublineages of the delta lineage carry this mutation, according to the Genomic surveillance circuit of Andalusia[29] (see http://nextstrain.clinbioinfosspa.es/SARS-COV-2-all?branchLabel=none&gt=N.377Y and Supplementary Figure S5).

Summarizing, the combined use of SARS-CoV-2 genome sequences and detailed clinical information of the patients allowed us to assess the impact of both, the SARS-CoV-2 lineage and the mutations each virus harbors, on mortality rate among patients hospitalized for COVID-19. These studies provide a more realistic and unbiased approach to define VOIs and VOCs.

## Supporting information

Suppementary Table S1

The Andalusian COVID-19 sequencing initiative

## Data Availability

Data are stored in the Base Poblacional de Salud (Population Health Database, see https://www.sspa.juntadeandalucia.es/servicioandaluzdesalud/profesionales/sistemas-de-informacion/base-poblacional-de-salud) and can be requested following the procedure for secondary use of clinical data approved by the Andalusian Ministry of Health (https://www.sspa.juntadeandalucia.es/servicioandaluzdesalud/sites/default/files/sincfiles/wsas-media-sas_normativa_mediafile/2021/resolucion_conjunta_acceso_a_datos_investigacion_def_20211201%28F%29.pdf)

## Acknowledgements

This research was funded by Spanish Ministry of Science and Innovation (grant PID2020-117979RB-I00), the Instituto de Salud Carlos III (ISCIII), co-funded with European Regional Development Funds (ERDF) (grant IMP/0019), and has also been funded by Consejería de Salud y Familias, Junta de Andalucía (grants COVID-0012-2020 and PS-2020-342) and the postdoctoral contract of Carlos Loucera (PAIDI2020-DOC_00350) co-funded by the European Social Fund (FSE) 2014-2020.

## Supplementary Material

**Supplementary Figure S1**, Circulation of the five SARS-CoV-2 variants eligible for the causal analysis in Andalusia (upper panel) and Spain (lower panel)

**Supplementary Figure S2**. Log Hazard ratios estimated for the 69 nucleotide mutations eligible for the causal analysis using the two approaches described in the text (the closed form estimator and the bootstrap). For each analysis an estimate of the LHR along with a 95% confidence interval and a p-value (FDR adjusted) of significance is provided.

**Supplementary Figure S3**. Correlations among the mutations in the SARS-CoV-2 genome significantly associated to patient survival.

**Supplementary figure S4**. Mutations occurring during the period studied (from 19th February 2020 to 30th April 2021) represented over the variants in which they appear in two phylogenetic formats. First column contains the mutation. Second column accounts for the evolutionary rates.. Third column with the time at which every variant was sampled from a patient.

**Supplementary Figure S5**. Presence of the mutation N:D377Y in the different SARS-CoV-2 viral genomes sampled in Andalusia according to the Genomic surveillance circuit of Andalusia. The upper branch corresponds to the delta variant and subtypes and the lower one to the almost extinct alpha variant.

See http://nextstrain.clinbioinfosspa.es/SARS-COV-2-ll?branchLabel=none&gt=N.377Y

**Supplementary Table S1**. ENA sample and project Ids of the SARS-CoV sequences used in this work

**Supplementary Table S2**. Nucleotide mutations eligible for causal analysis. The first column is the mutation name; the second is the position; the third column, labeled as CDS, is the protein affected; the fourth column is the amino acid mutation name; the fifth column is the number of variants bearing this mutation; and the following columns provide the values of the two approaches for hazard ratio estimation, the closed form, with the hazard ratio coefficient, SD, confidence intervals 5 and 95, the p-value and the FDR adjusted p-value, and the bootstrap approach with the HR coefficients (Boot. Statistic), bias, SD, confidence intervals 5 and 95 and the last column, labeled as Boot, indicates if significance is confirmed by bootstrap (T: true and F: false)

## References

1. Shu Y, McCauley J. GISAID: Global initiative on sharing all influenza data– from vision to reality. Eurosurveillance. 2017;22(13):30494.

2. Faria NR, Mellan TA, Whittaker C, et al. Genomics and epidemiology of the P. 1 SARS-CoV-2 lineage in Manaus, Brazil. Science. 2021;372(6544):815–821.

3. Tang JW, Tambyah PA, Hui DS. Emergence of a new SARS-CoV-2 variant in the UK. Journal of Infection. 2021;82(4):e27–e28.

4. Tegally H, Wilkinson E, Giovanetti M, et al. Detection of a SARS-CoV-2 variant of concern in South Africa. Nature. 2021;592(7854):438–443.

5. Volz E, Mishra S, Chand M, et al. Assessing transmissibility of SARS-CoV-2 lineage B. 1.1. 7 in England. Nature. 2021;593(7858):266–269.

6. Hodcroft EB, Zuber M, Nadeau S, et al. Spread of a SARS-CoV-2 variant through Europe in the summer of 2020. Nature. 2021;595(7869):707–712.

7. Zhou D, Dejnirattisai W, Supasa P, et al. Evidence of escape of SARS-CoV-2 variant B. 1.351 from natural and vaccine-induced sera. Cell. 2021;184(9):2348-2361. e6.

8. Chen RE, Zhang X, Case JB, et al. Resistance of SARS-CoV-2 variants to neutralization by monoclonal and serum-derived polyclonal antibodies. Nature medicine. 2021;27(4):717–726.

9. Cyranoski D. Alarming COVID variants show vital role of genomic surveillance. Nature. 2021;589(7842).

10. WHO. SARS-CoV-2 variants of concern and variants of interest. World Health Organization 2021 [cited 2021 21/09/]. Available from: https://www.who.int/en/activities/tracking-SARS-CoV-2-variants/

11. Kwok AJ, Mentzer A, Knight JC. Host genetics and infectious disease: new tools, insights and translational opportunities. Nature Reviews Genetics. 2021;22(3):137–153.

12. Fallerini C, Daga S, Mantovani S, et al. Association of Toll-like receptor 7 variants with life-threatening COVID-19 disease in males: findings from a nested case-control study. eLife. 2021;10:e67569.

13. Severe_Covid-19_GWAS_Group. Genomewide association study of severe Covid-19 with respiratory failure. New England Journal of Medicine. 2020;383(16):1522–1534.

14. Zhou S, Butler-Laporte G, Nakanishi T, et al. A Neanderthal OAS1 isoform protects individuals of European ancestry against COVID-19 susceptibility and severity. Nature medicine. 2021;27(4):659–667.

15. COVID-19_Host_Genetics_Initiative. Mapping the human genetic architecture of COVID-19 by worldwide meta-analysis. MedRxiv. 2021.

16. Kousathanas A, Pairo-Castineira E, Rawlik K, et al. Whole genome sequencing identifies multiple loci for critical illness caused by COVID-19. medRxiv. 2021:2021.09.02.21262965.

17. Davies NG, Jarvis CI, Edmunds WJ, et al. Increased mortality in community-tested cases of SARS-CoV-2 lineage B. 1.1. 7. Nature. 2021;593(7858):270–274.

18. Davies NG, Abbott S, Barnard RC, et al. Estimated transmissibility and impact of SARS-CoV-2 lineage B. 1.1. 7 in England. Science. 2021.

19. Frampton D, Rampling T, Cross A, et al. Genomic characteristics and clinical effect of the emergent SARS-CoV-2 B. 1.1. 7 lineage in London, UK: a whole-genome sequencing and hospital-based cohort study. The Lancet infectious diseases. 2021.

20. Toyoshima Y, Nemoto K, Matsumoto S, et al. SARS-CoV-2 genomic variations associated with mortality rate of COVID-19. Journal of human genetics. 2020;65(12):1075–1082.

21. Nagy Á, Pongor S, Győrffy B. Different mutations in SARS-CoV-2 associate with severe and mild outcome. International journal of antimicrobial agents. 2021;57(2):106272.

22. Young BE, Fong S-W, Chan Y-H, et al. Effects of a major deletion in the SARS-CoV-2 genome on the severity of infection and the inflammatory response: an observational cohort study. The Lancet. 2020;396(10251):603–611.

23. CDC. SARS-CoV-2 variant classifications and definitions. Centers for Disease Control and Prevention 2021 [cited 2021]. Available from: https://www.cdc.gov/coronavirus/2019-ncov/variants/variant-info.html

24. The_COVID-19_Genomics_UK_(COG-UK)_consortium. An integrated national scale SARS-CoV-2 genomic surveillance network. The Lancet Microbe. 2020;1(3):e99.

25. Tao K, Tzou PL, Nouhin J, et al. The biological and clinical significance of emerging SARS-CoV-2 variants. Nature Reviews Genetics. 2021:1–17.

26. Outbreak.info. A standardized, open-source database of COVID-19 resources and epidemiology data 2021. Available from: https://outbreak.info/

27. Sequencing of the SARS-CoV-2 virus genome for the monitoring and management of the Covid-19 epidemic in Andalusia and the rapid generation of prognostic and response to treatment biomarkers 2020 [cited 2021 22/09/]. Available from: https://www.clinbioinfosspa.es/projects/covseq/indexEng.html

28. SARS-CoV-2 whole genome sequencing circuit in Andalusia 2021. Available from: https://www.clinbioinfosspa.es/COVID_circuit/

29. Dopazo J, Maya-Miles D, García F, et al. Implementing Personalized Medicine in COVID-19 in Andalusia: An Opportunity to Transform the Healthcare System. Journal of Personalized Medicine. 2021;11(6):475.

30. Muñoyerro-Muñiz D, Goicoechea-Salazar J, García-León F, et al. Health record linkage: Andalusian health population database. Gaceta Sanitaria. 2019;34(2):105–113.

31. SeqCOVID, genomic epidemiology of SARS-CoV-2 in Spain 2020 [cited 2022 21/01/]. Available from: https://seqcovid.csic.es/

32. ISCIII. Integration of genome sequencing in the SARS-CoV-2 surveillance (in Spanish) 2021. Available from: https://www.mscbs.gob.es/profesionales/saludPublica/ccayes/alertasActual/nCov/documentos/Integracion_de_la_secuenciacion_genomica-en_la_vigilancia_del_SARS-CoV-2.pdf

33. Artic_Network. SARS-CoV-2 amplicon set V3 2021. Available from: https://artic.network/ncov-2019

34. Patel H, Varona S, Monzón S, et al. Nf-Core/Viral recon v1.1 - Steel Pangolin (Version 1.1.0) 2021. Available from: https://zenodo.org/record/3905178#.YW2i9Joza70

35. Wu F, Zhao S, Yu B, et al. A new coronavirus associated with human respiratory disease in China. Nature. 2020;579(7798):265–269.

36. Langmead B, Salzberg SL. Fast gapped-read alignment with Bowtie 2. Nat Methods. 2012 Apr;9(4):357–9.

37. Grubaugh ND, Gangavarapu K, Quick J, et al. An amplicon-based sequencing framework for accurately measuring intrahost virus diversity using PrimalSeq and iVar. Genome biology. 2019;20(1):1–19.

38. Picard_team. Picard. A set of command line tools (in Java) for manipulating high-throughput sequencing (HTS) data and formats such as SAM/BAM/CRAM and VCF. 2014.

39. Aksamentov I, Neher RA. Nextclade. Viral genome clade assignment, mutation calling, and sequence quality checks 2020. Available from: https://github.com/nextstrain/nextclade

40. O’Toole Á, Scher E, Underwood A, et al. Assignment of epidemiological lineages in an emerging pandemic using the pangolin tool. Virus Evolution. 2021;7(2):veab064.

41. Huddleston J, Hadfield J, Sibley TR, et al. Augur: a bioinformatics toolkit for phylogenetic analyses of human pathogens. Journal of Open Source Software. 2021;6(57):2906.

42. Minh BQ, Schmidt HA, Chernomor O, et al. IQ-TREE 2: New models and efficient methods for phylogenetic inference in the genomic era. Molecular biology and evolution. 2020;37(5):1530–1534.

43. Tavaré S. Some probabilistic and statistical problems in the analysis of DNA sequences. Lectures on mathematics in the life sciences. 1986;17(2):57–86.

44. Infrastructure for secure generation of Real World Evidence from Real World Data from the Andalusian Health Population Database 2020 [cited 2021 17/11/]. Available from: https://www.clinbioinfosspa.es/projects/iRWD/

45. Sterne JA, Murthy S, Diaz JV, et al. Association between administration of systemic corticosteroids and mortality among critically ill patients with COVID-19: a meta-analysis. JAMA : the journal of the American Medical Association. 2020;324(13):1330–1341.

46. Stuart EA, Lee BK, Leacy FP. Prognostic score–based balance measures can be a useful diagnostic for propensity score methods in comparative effectiveness research. Journal of clinical epidemiology. 2013;66(8):S84-S90. e1.

47. Hajage D, Chauvet G, Belin L, et al. Closed-form variance estimator for weighted propensity score estimators with survival outcome. Biometrical Journal. 2018;60(6):1151–1163.

48. Austin PC. Variance estimation when using inverse probability of treatment weighting (IPTW) with survival analysis. Statistics in medicine. 2016;35(30):5642–5655.

49. Greifer N. WeightIt: Weighting for Covariate Balance in Observational Studies 2021. Available from: https://cran.r-project.org/package=WeightIt

50. Greifer N. cobalt: Covariate Balance Tables and Plots 2021. Available from: https://cran.r-project.org/package=cobalt

51. Gutiérrez-Gutiérrez B, del Toro MD, Borobia AM, et al. Identification and validation of clinical phenotypes with prognostic implications in patients admitted to hospital with COVID-19: a multicentre cohort study. The Lancet infectious diseases. 2021 2021/06/01/;21(6):783–792.

52. Benjamini Y, Yekutieli D. The control of false discovery rate in multiple testing under dependency. Annals of Statistics. 2001;29:1165–1188.

53. CoVariants 2020 [cited 2021 25/03/]. Available from: https://covariants.org/

54. Elbe S, Buckland-Merrett G. Data, disease and diplomacy: GISAID’s innovative contribution to global health. Glob Chall. 2017 Jan;1(1):33–46.

55. Rambaut A, Holmes EC, O’Toole Á, et al. A dynamic nomenclature proposal for SARS-CoV-2 lineages to assist genomic epidemiology. Nature microbiology. 2020;5(11):1403–1407.

56. Mistry J, Chuguransky S, Williams L, et al. Pfam: The protein families database in 2021. Nucleic Acids Res. 2021 Jan 8;49(D1):D412–d419.

57. Lu G, Wang Q, Gao GF. Bat-to-human: spike features determining ‘host jump’ of coronaviruses SARS-CoV, MERS-CoV, and beyond. Trends Microbiol. 2015 Aug;23(8):468–78.

58. Flower TG, Buffalo CZ, Hooy RM, et al. Structure of SARS-CoV-2 ORF8, a rapidly evolving immune evasion protein. Proceedings of the National Academy of Sciences. 2021;118(2).

59. Tan Y, Schneider T, Leong M, et al. Novel Immunoglobulin Domain Proteins Provide Insights into Evolution and Pathogenesis of SARS-CoV-2-Related Viruses. mBio. 2020 May 29;11(3).

60. Graham RL, Baric RS. Recombination, reservoirs, and the modular spike: mechanisms of coronavirus cross-species transmission. J Virol. 2010 Apr;84(7):3134–46.

61. Candido DS, Claro IM, De Jesus JG, et al. Evolution and epidemic spread of SARS-CoV-2 in Brazil. Science. 2020;369(6508):1255–1260.

62. Tegally H, Wilkinson E, Lessells RJ, et al. Sixteen novel lineages of SARS-CoV-2 in South Africa. Nature Medicine. 2021;27(3):440–446.

63. Gangavarapu K, Latiff AA, Mullen JL, et al. Outbreak. info genomic reports: scalable and dynamic surveillance of SARS-CoV-2 variants and mutations. medRxiv. 2022.

64. Chen C, Nadeau S, Yared M, et al. CoV-Spectrum: analysis of globally shared SARS-CoV-2 data to identify and characterize new variants. Bioinformatics. 2022;38(6):1735–1737.

