## Supplementary Figure S1 for "Assessing the impact of SARS-CoV-2 lineages and mutations on patient survival"

Circulation of the five SARS-CoV-2 variants eligible for the causal analysis in A) Andalusia, B) Spain, C) France, D) United Kingdom, and E) Portugal.

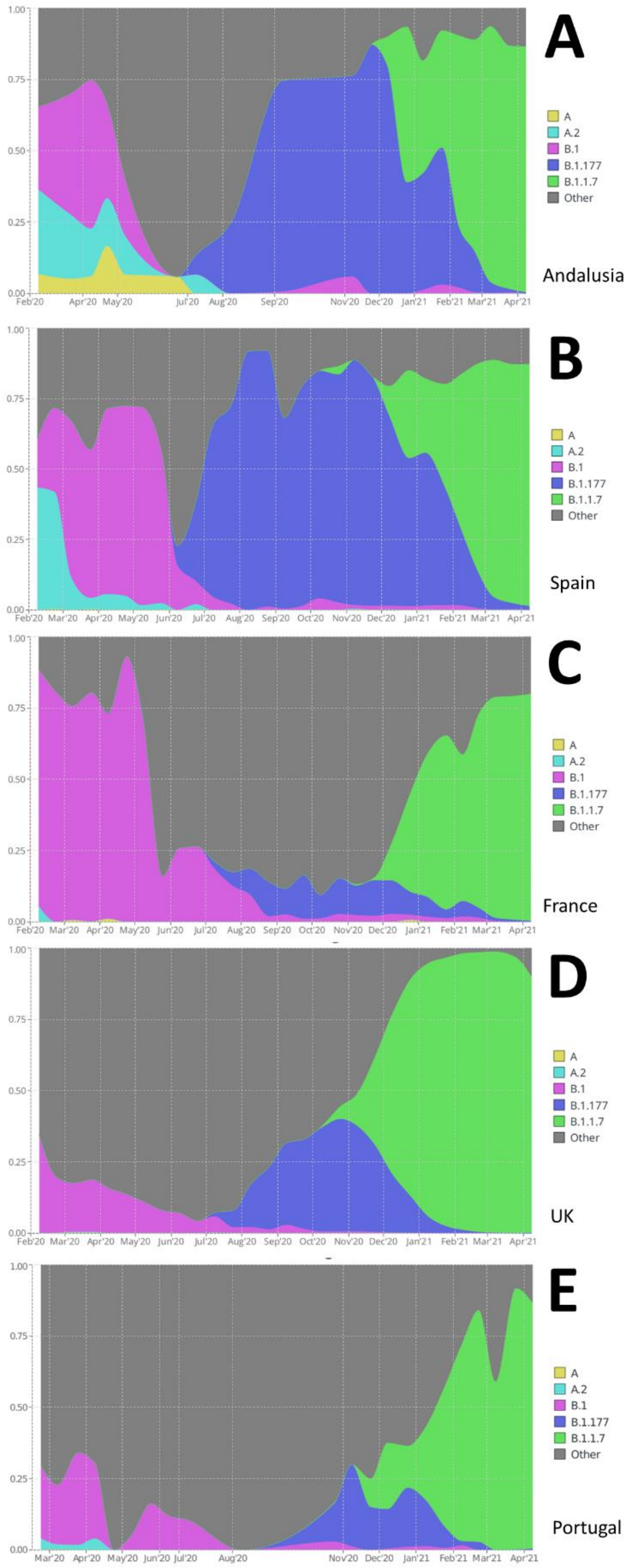
