## Supplementary Figure S2 for "Assessing the impact of SARS-CoV-2 lineages and mutations on patient survival"

Log Hazard ratios estimated for the 69 nucleotide mutations eligible for the causal analysis using the two approaches described in the text (the closed form estimator and the bootstrap). For each analysis an estimate of the LHR along with a 95% confidence interval and a p-value (FDR adjusted) of significance is provided.

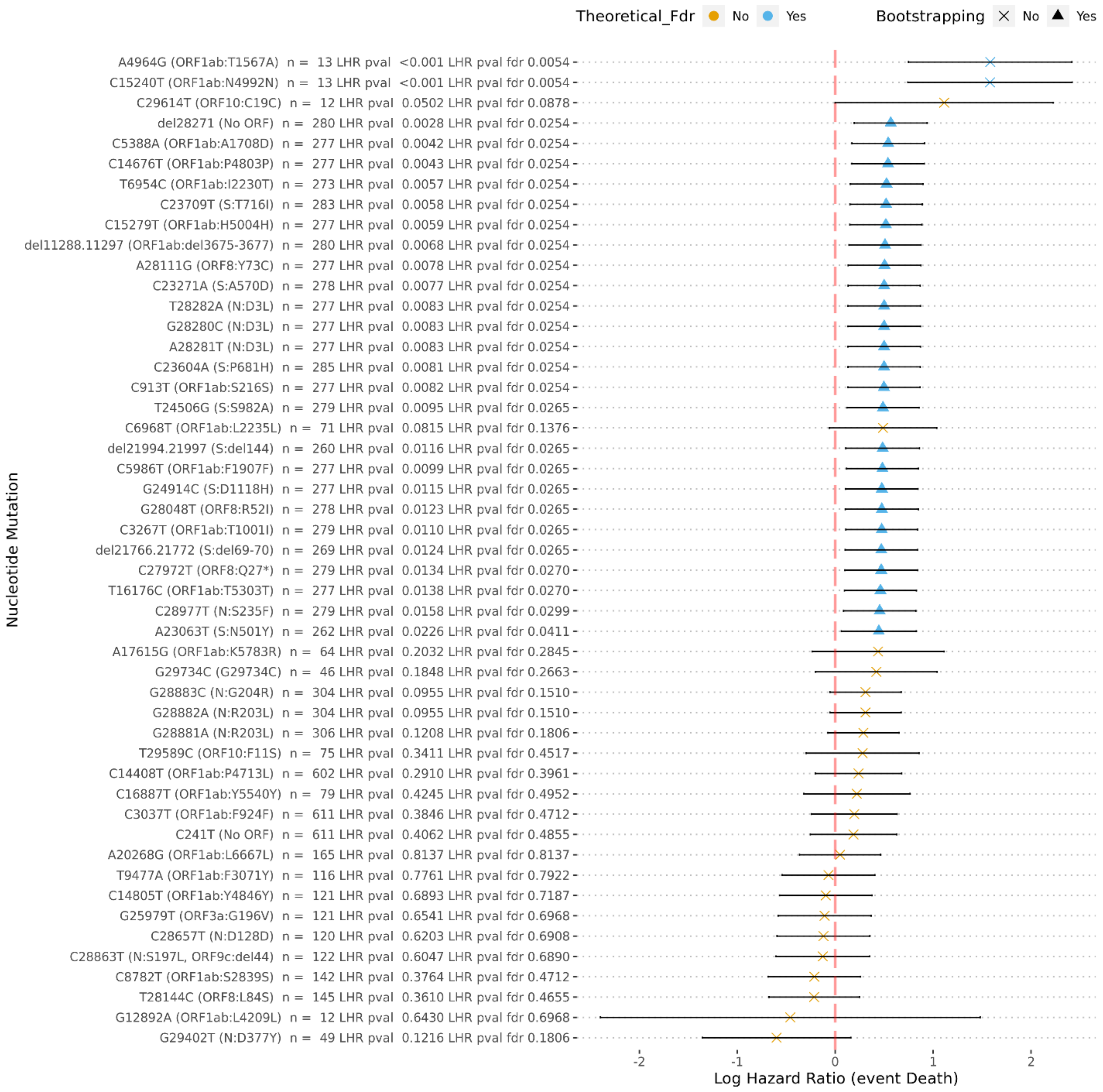
