## Supplementary Figure S3 for "Assessing the impact of SARS-CoV-2 lineages and mutations on patient survival"

Correlations among the mutations in the SARS-CoV-2 genome significantly associated to patient survival.

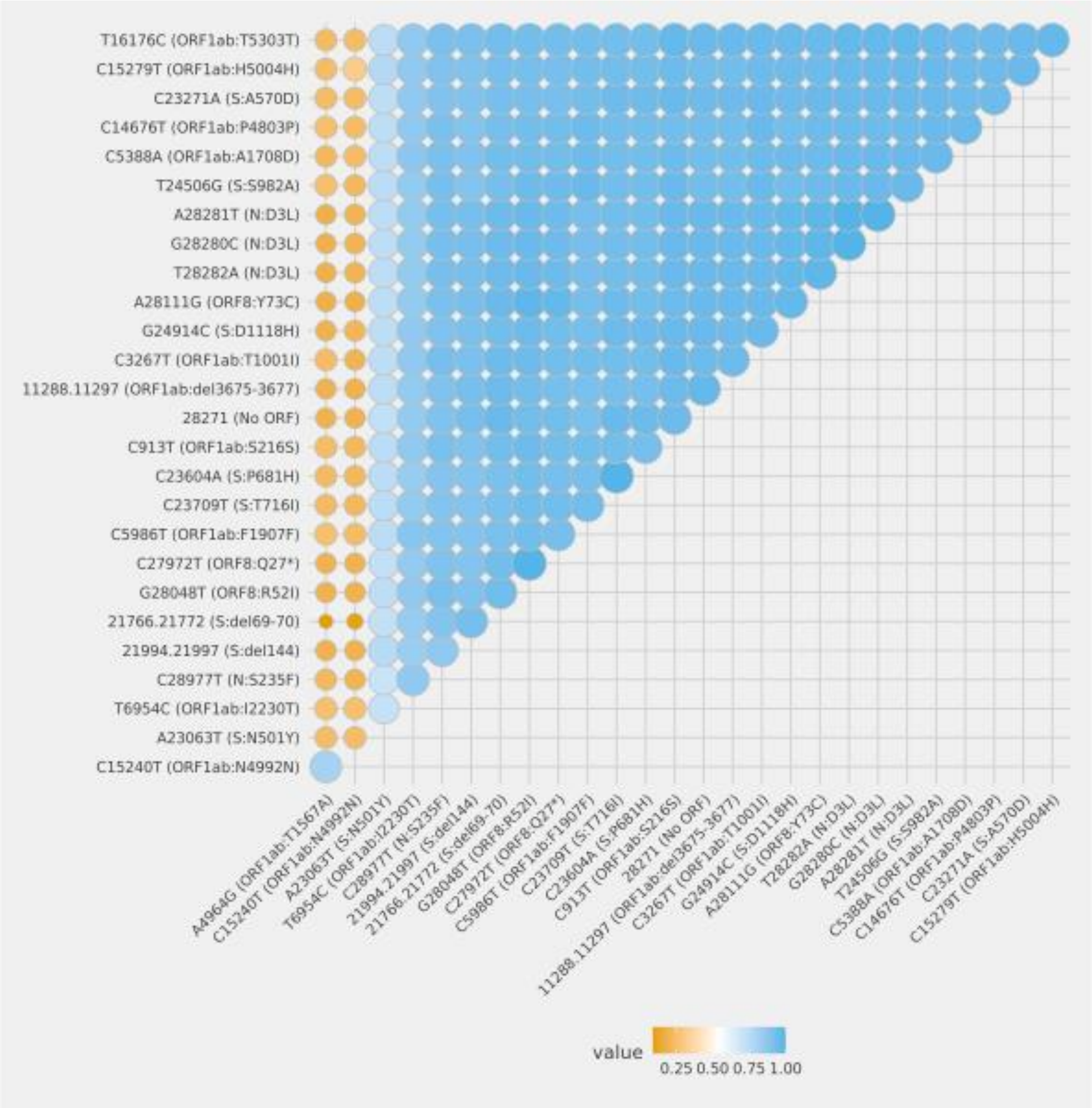
