## Supplementary Figure S4 for "Assessing the impact of SARS-CoV-2 lineages and mutations on patient survival"

Mutations occurring during the period studied (from 19th February 2020 to 30th April 2021) represented over the variants in which they appear in two phylogenetic formats. First column contains the mutation. Second column accounts for the evolutionary rates. Third column with the time at which every variant was sampled from a patient.

**Mutation** | **Clock-like phylogeny** | **Phylogeny with time in the X axis**

ORF1ab: A1708D

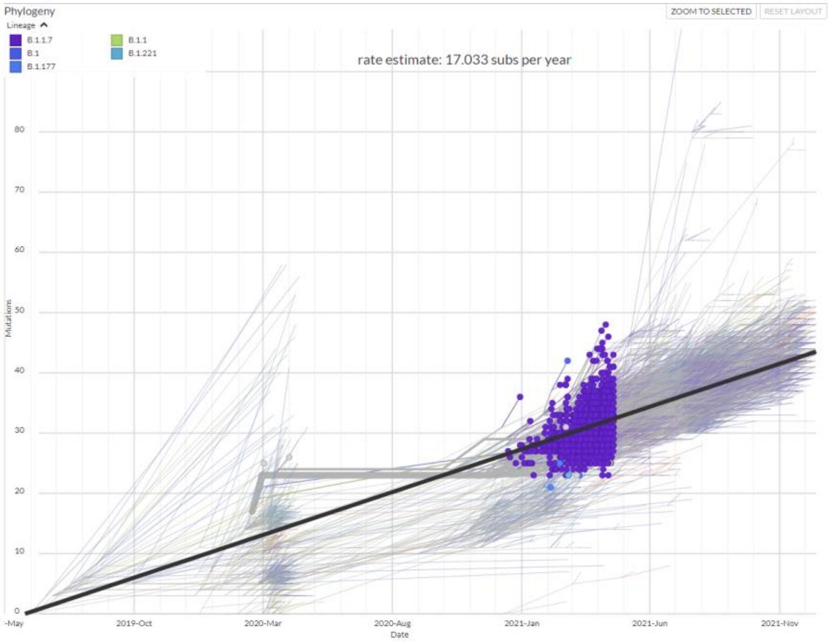


**A**

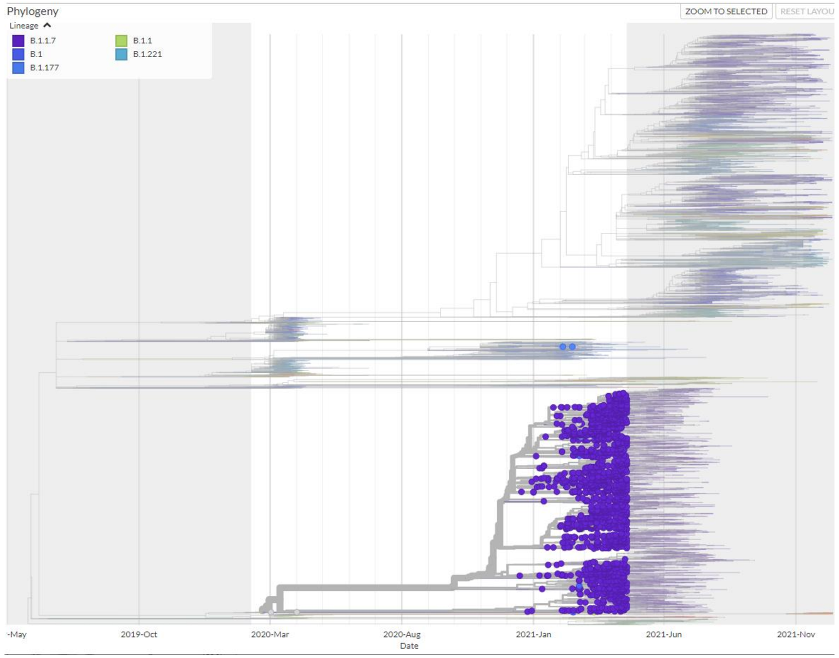


**B**

ORF1ab:T1567A

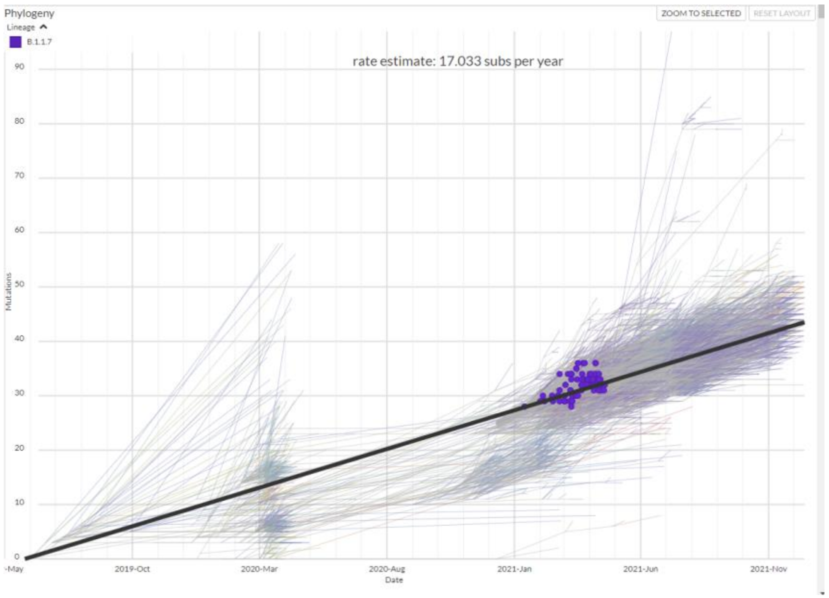


C

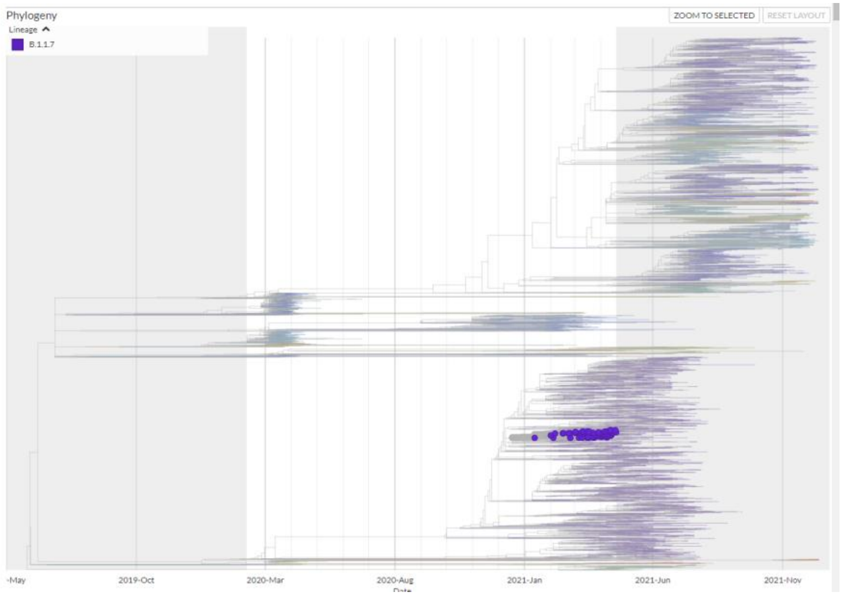


D

N:D377Y

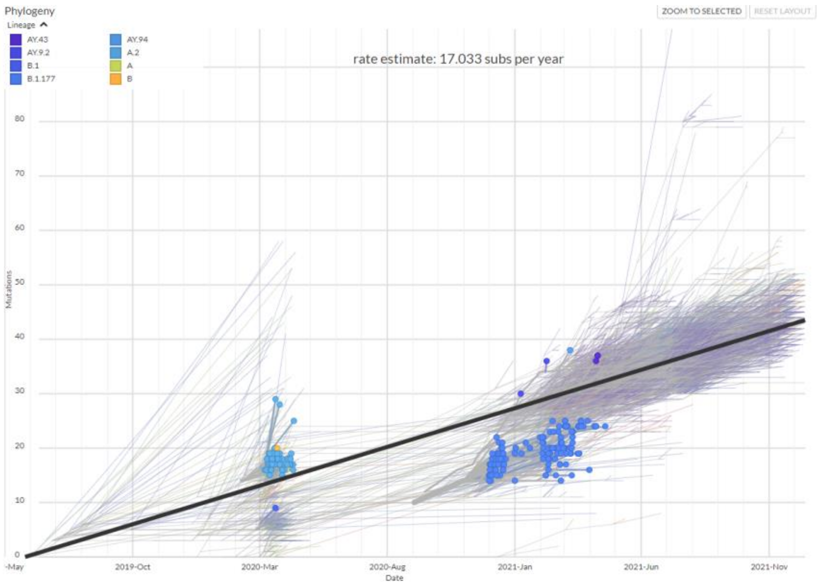


E

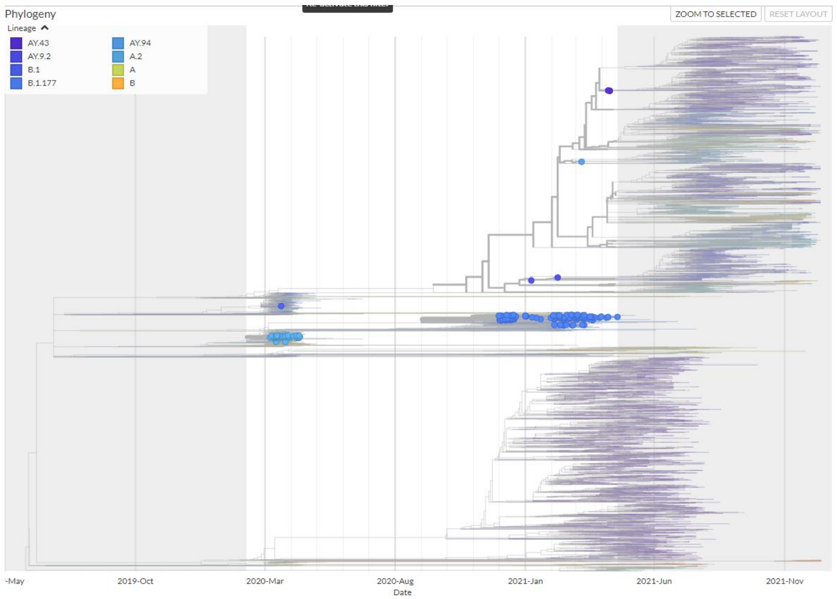


F
