## Supplementary Figure S5 for "Assessing the impact of SARS-CoV-2 lineages and mutations on patient survival"

Presence of the mutation N:D377Y in the different SARS-CoV-2 viral genomes sampled in Andalusia according to the Genomic surveillance circuit of Andalusia. The upper branch corresponds to the delta variant and subtypes and the lower one to the almost extinct alpha variant.

See http://nextstrain.clinbioinfosspa.es/SARS-COV-2-all?branchLabel=none&gt=N.377Y

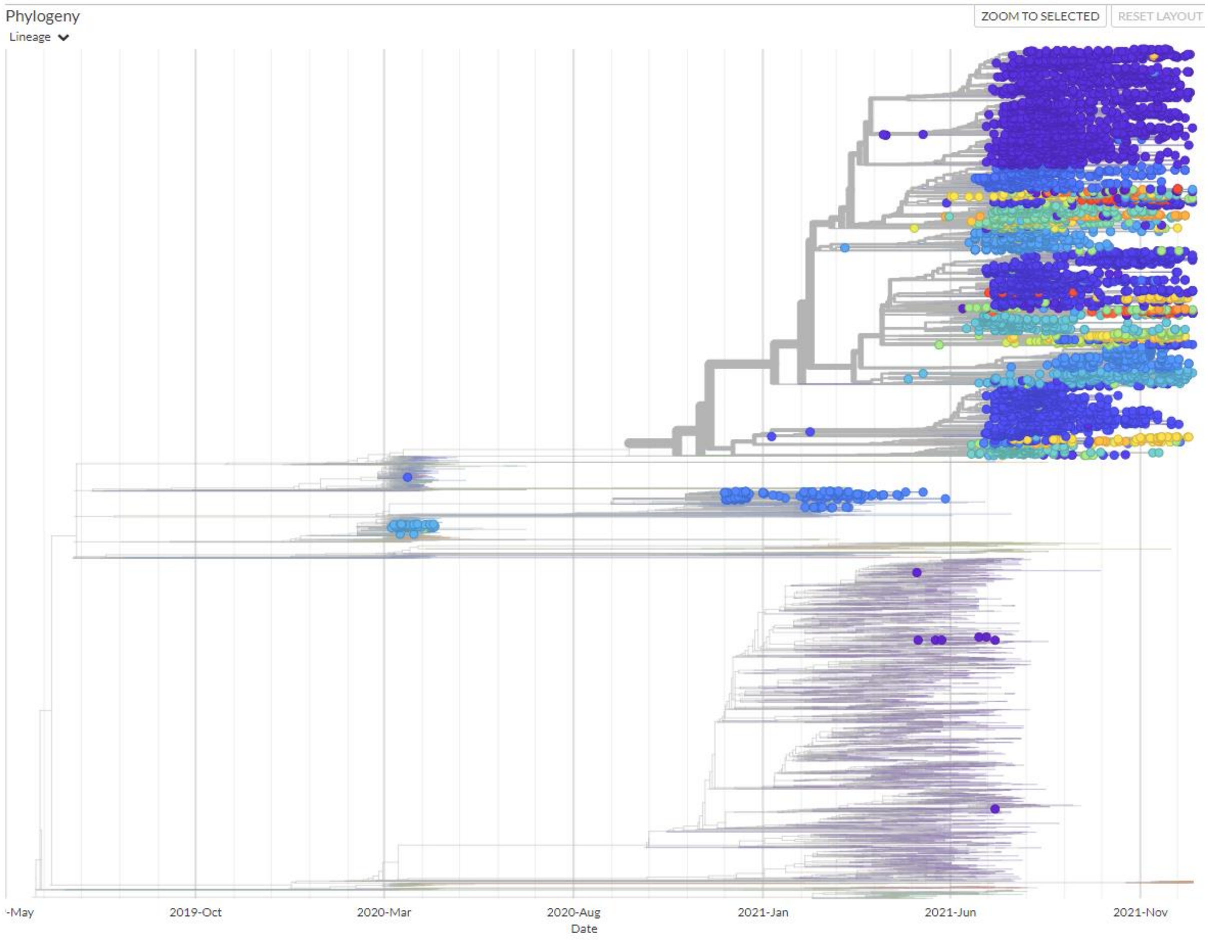
