## Supplementary material for "Assessing the impact of SARS-CoV-2 lineages and mutations on patient survival": Suppementary Table S1

### Supplementary Table S1.

ENA sample and project Ids of the SARS-CoV sequences used in this work

| ENA sample ID | ENA project ID |
|---|---|
| SAMEA10306558 | PRJEB44396 |
| SAMEA10305326 | PRJEB44396 |
| SAMEA10305325 | PRJEB44396 |
| SAMEA10305315 | PRJEB44396 |
| SAMEA10305310 | PRJEB44396 |
| SAMEA10305307 | PRJEB44396 |
| SAMEA10305297 | PRJEB44396 |
| SAMEA10305293 | PRJEB44396 |
| SAMEA10304167 | PRJEB44396 |
| SAMEA10304151 | PRJEB44396 |
| SAMEA10304142 | PRJEB44396 |
| SAMEA10304135 | PRJEB44396 |
| SAMEA10304117 | PRJEB44396 |
| SAMEA10304111 | PRJEB44396 |
| SAMEA10304110 | PRJEB44396 |
| SAMEA10304108 | PRJEB44396 |
| SAMEA10304104 | PRJEB44396 |
| SAMEA10304101 | PRJEB44396 |
| SAMEA10304096 | PRJEB44396 |
| SAMEA10304090 | PRJEB44396 |
| SAMEA10304085 | PRJEB44396 |
| SAMEA10304083 | PRJEB44396 |
| SAMEA10304079 | PRJEB44396 |
| SAMEA10304078 | PRJEB44396 |
| SAMEA10304077 | PRJEB44396 |
| SAMEA10304076 | PRJEB44396 |
| SAMEA10304075 | PRJEB44396 |
| SAMEA10304074 | PRJEB44396 |
| SAMEA10304069 | PRJEB44396 |
| SAMEA10304068 | PRJEB44396 |
| SAMEA10304058 | PRJEB44396 |
| SAMEA10304056 | PRJEB44396 |
| SAMEA10304054 | PRJEB44396 |
| SAMEA10304049 | PRJEB44396 |
| SAMEA10304045 | PRJEB44396 |
| SAMEA10304043 | PRJEB44396 |
| SAMEA10304042 | PRJEB44396 |
| SAMEA10299902 | PRJEB44396 |
| SAMEA10299886 | PRJEB44396 |
| SAMEA10299795 | PRJEB44396 |
| SAMEA10299749 | PRJEB44396 |
| SAMEA10299528 | PRJEB44396 |

| | |
|---|---|
| SAMEA10299520 | PRJEB44396 |
| SAMEA10299515 | PRJEB44396 |
| SAMEA10299508 | PRJEB44396 |
| SAMEA10299504 | PRJEB44396 |
| SAMEA10299503 | PRJEB44396 |
| SAMEA10299478 | PRJEB44396 |
| SAMEA8729476 | PRJEB44396 |
| SAMEA8722672 | PRJEB44396 |
| SAMEA8722663 | PRJEB44396 |
| SAMEA8693734 | PRJEB44396 |
| SAMEA8693691 | PRJEB44396 |
| SAMEA8693687 | PRJEB44396 |
| SAMEA8693686 | PRJEB44396 |
| SAMEA8693373 | PRJEB44396 |
| SAMEA8693372 | PRJEB44396 |
| SAMEA8693366 | PRJEB44396 |
| SAMEA8693364 | PRJEB44396 |
| SAMEA8693359 | PRJEB44396 |
| SAMEA8693341 | PRJEB44396 |
| SAMEA8693324 | PRJEB44396 |
| SAMEA8693321 | PRJEB44396 |
| SAMEA8693310 | PRJEB44396 |
| SAMEA8693307 | PRJEB44396 |
| SAMEA8693301 | PRJEB44396 |
| SAMEA8693281 | PRJEB44396 |
| SAMEA8693279 | PRJEB44396 |
| SAMEA8693273 | PRJEB44396 |
| SAMEA8693269 | PRJEB44396 |
| SAMEA8693263 | PRJEB44396 |
| SAMEA8693261 | PRJEB44396 |
| SAMEA8693255 | PRJEB44396 |
| SAMEA8694675 | PRJEB44396 |
| SAMEA8694671 | PRJEB44396 |
| SAMEA8694660 | PRJEB44396 |
| SAMEA8694650 | PRJEB44396 |
| SAMEA8694642 | PRJEB44396 |
| SAMEA8694632 | PRJEB44396 |
| SAMEA8694629 | PRJEB44396 |
| SAMEA8694624 | PRJEB44396 |
| SAMEA8694617 | PRJEB44396 |
| SAMEA8694613 | PRJEB44396 |
| SAMEA8694602 | PRJEB44396 |
| SAMEA8694600 | PRJEB44396 |
| SAMEA8694592 | PRJEB44396 |
| SAMEA8694589 | PRJEB44396 |
| SAMEA8694587 | PRJEB44396 |
| SAMEA8694575 | PRJEB44396 |
| SAMEA8694571 | PRJEB44396 |

| | |
|---|---|
| SAMEA8694567 | PRJEB44396 |
| SAMEA8694566 | PRJEB44396 |
| SAMEA8694554 | PRJEB44396 |
| SAMEA8694547 | PRJEB44396 |
| SAMEA8694542 | PRJEB44396 |
| SAMEA8694540 | PRJEB44396 |
| SAMEA8694534 | PRJEB44396 |
| SAMEA8694530 | PRJEB44396 |
| SAMEA8694524 | PRJEB44396 |
| SAMEA8694523 | PRJEB44396 |
| SAMEA8694517 | PRJEB44396 |
| SAMEA8697543 | PRJEB44396 |
| SAMEA8697541 | PRJEB44396 |
| SAMEA8697539 | PRJEB44396 |
| SAMEA8697536 | PRJEB44396 |
| SAMEA8697528 | PRJEB44396 |
| SAMEA8697524 | PRJEB44396 |
| SAMEA8697515 | PRJEB44396 |
| SAMEA8697514 | PRJEB44396 |
| SAMEA8697512 | PRJEB44396 |
| SAMEA8697501 | PRJEB44396 |
| SAMEA8697496 | PRJEB44396 |
| SAMEA8697485 | PRJEB44396 |
| SAMEA8697478 | PRJEB44396 |
| SAMEA8697475 | PRJEB44396 |
| SAMEA8697471 | PRJEB44396 |
| SAMEA8697470 | PRJEB44396 |
| SAMEA8697469 | PRJEB44396 |
| SAMEA8697468 | PRJEB44396 |
| SAMEA8697465 | PRJEB44396 |
| SAMEA8697456 | PRJEB44396 |
| SAMEA8697436 | PRJEB44396 |
| SAMEA8697431 | PRJEB44396 |
| SAMEA8697430 | PRJEB44396 |
| SAMEA8697428 | PRJEB44396 |
| SAMEA8697421 | PRJEB44396 |
| SAMEA8697395 | PRJEB44396 |
| SAMEA8697393 | PRJEB44396 |
| SAMEA8697391 | PRJEB44396 |
| SAMEA8697381 | PRJEB44396 |
| SAMEA8697380 | PRJEB44396 |
| SAMEA8697379 | PRJEB44396 |
| SAMEA8697377 | PRJEB44396 |
| SAMEA8710066 | PRJEB44396 |
| SAMEA8710062 | PRJEB44396 |
| SAMEA8710061 | PRJEB44396 |
| SAMEA8710058 | PRJEB44396 |
| SAMEA8710052 | PRJEB44396 |

| | |
|---|---|
| SAMEA8710051 | PRJEB44396 |
| SAMEA8710042 | PRJEB44396 |
| SAMEA8710040 | PRJEB44396 |
| SAMEA8710035 | PRJEB44396 |
| SAMEA8710033 | PRJEB44396 |
| SAMEA8710030 | PRJEB44396 |
| SAMEA8710025 | PRJEB44396 |
| SAMEA8710011 | PRJEB44396 |
| SAMEA8709999 | PRJEB44396 |
| SAMEA8709995 | PRJEB44396 |
| SAMEA8709987 | PRJEB44396 |
| SAMEA8709965 | PRJEB44396 |
| SAMEA8709955 | PRJEB44396 |
| SAMEA8709947 | PRJEB44396 |
| SAMEA8709943 | PRJEB44396 |
| SAMEA8709938 | PRJEB44396 |
| SAMEA8709933 | PRJEB44396 |
| SAMEA8709930 | PRJEB44396 |
| SAMEA10299437 | PRJEB44396 |
| SAMEA10299436 | PRJEB44396 |
| SAMEA10299432 | PRJEB44396 |
| SAMEA10299426 | PRJEB44396 |
| SAMEA10299404 | PRJEB44396 |
| SAMEA10299402 | PRJEB44396 |
| SAMEA10299397 | PRJEB44396 |
| SAMEA10299386 | PRJEB44396 |
| SAMEA10299385 | PRJEB44396 |
| SAMEA8717253 | PRJEB44396 |
| SAMEA8717245 | PRJEB44396 |
| SAMEA8717244 | PRJEB44396 |
| SAMEA8717243 | PRJEB44396 |
| SAMEA8717237 | PRJEB44396 |
| SAMEA8717233 | PRJEB44396 |
| SAMEA8717226 | PRJEB44396 |
| SAMEA8717217 | PRJEB44396 |
| SAMEA8717211 | PRJEB44396 |
| SAMEA8717207 | PRJEB44396 |
| SAMEA8717189 | PRJEB44396 |
| SAMEA8717178 | PRJEB44396 |
| SAMEA8717164 | PRJEB44396 |
| SAMEA8717160 | PRJEB44396 |
| SAMEA8717157 | PRJEB44396 |
| SAMEA8717155 | PRJEB44396 |
| SAMEA8717151 | PRJEB44396 |
| SAMEA8717150 | PRJEB44396 |
| SAMEA8717136 | PRJEB44396 |
| SAMEA8717135 | PRJEB44396 |
| SAMEA8717129 | PRJEB44396 |

| | |
|---|---|
| SAMEA8717128 | PRJEB44396 |
| SAMEA8717123 | PRJEB44396 |
| SAMEA8712601 | PRJEB44396 |
| SAMEA8712588 | PRJEB44396 |
| SAMEA10299370 | PRJEB44396 |
| SAMEA10299369 | PRJEB44396 |
| SAMEA10299365 | PRJEB44396 |
| SAMEA10299357 | PRJEB44396 |
| SAMEA10299354 | PRJEB44396 |
| SAMEA10299349 | PRJEB44396 |
| SAMEA10299346 | PRJEB44396 |
| SAMEA10299339 | PRJEB44396 |
| SAMEA10299337 | PRJEB44396 |
| SAMEA10299334 | PRJEB44396 |
| SAMEA10299330 | PRJEB44396 |
| SAMEA10299326 | PRJEB44396 |
| SAMEA10299325 | PRJEB44396 |
| SAMEA10299320 | PRJEB44396 |
| SAMEA10299312 | PRJEB44396 |
| SAMEA10299307 | PRJEB44396 |
| SAMEA10299305 | PRJEB44396 |
| SAMEA10299303 | PRJEB44396 |
| SAMEA8724636 | PRJEB44396 |
| SAMEA8724634 | PRJEB44396 |
| SAMEA8724627 | PRJEB44396 |
| SAMEA8724625 | PRJEB44396 |
| SAMEA8724618 | PRJEB44396 |
| SAMEA8724602 | PRJEB44396 |
| SAMEA8724583 | PRJEB44396 |
| SAMEA8724572 | PRJEB44396 |
| SAMEA8724564 | PRJEB44396 |
| SAMEA8724563 | PRJEB44396 |
| SAMEA8724559 | PRJEB44396 |
| SAMEA8724555 | PRJEB44396 |
| SAMEA8724542 | PRJEB44396 |
| SAMEA8724536 | PRJEB44396 |
| SAMEA8724533 | PRJEB44396 |
| SAMEA8724526 | PRJEB44396 |
| SAMEA8724520 | PRJEB44396 |
| SAMEA8724519 | PRJEB44396 |
| SAMEA8724514 | PRJEB44396 |
| SAMEA8724513 | PRJEB44396 |
| SAMEA8724506 | PRJEB44396 |
| SAMEA10299300 | PRJEB44396 |
| SAMEA10299282 | PRJEB44396 |
| SAMEA10299259 | PRJEB44396 |
| SAMEA10299250 | PRJEB44396 |
| SAMEA10299244 | PRJEB44396 |

| | |
|---|---|
| SAMEA10299229 | PRJEB44396 |
| SAMEA10299223 | PRJEB44396 |
| SAMEA10291327 | PRJEB44396 |
| SAMEA10291238 | PRJEB44396 |
| SAMEA10291350 | PRJEB44396 |
| SAMEA10291333 | PRJEB44396 |
| SAMEA10291353 | PRJEB44396 |
| SAMEA10291326 | PRJEB44396 |
| SAMEA10291292 | PRJEB44396 |
| SAMEA10291323 | PRJEB44396 |
| SAMEA10291231 | PRJEB44396 |
| SAMEA10291343 | PRJEB44396 |
| SAMEA10291336 | PRJEB44396 |
| SAMEA10291281 | PRJEB44396 |
| SAMEA10291273 | PRJEB44396 |
| SAMEA10291227 | PRJEB44396 |
| SAMEA10291372 | PRJEB44396 |
| SAMEA10291369 | PRJEB44396 |
| SAMEA10291311 | PRJEB44396 |
| SAMEA10291263 | PRJEB44396 |
| SAMEA10291223 | PRJEB44396 |
| SAMEA10291254 | PRJEB44396 |
| SAMEA10291341 | PRJEB44396 |
| SAMEA10291247 | PRJEB44396 |
| SAMEA10291306 | PRJEB44396 |
| SAMEA10291214 | PRJEB44396 |
| SAMEA10291244 | PRJEB44396 |
| SAMEA10291340 | PRJEB44396 |
| SAMEA10291211 | PRJEB44396 |
| SAMEA10291347 | PRJEB44396 |
| SAMEA10291144 | PRJEB44396 |
| SAMEA10291140 | PRJEB44396 |
| SAMEA10291190 | PRJEB44396 |
| SAMEA10291160 | PRJEB44396 |
| SAMEA10291134 | PRJEB44396 |
| SAMEA10291133 | PRJEB44396 |
| SAMEA10291195 | PRJEB44396 |
| SAMEA10291131 | PRJEB44396 |
| SAMEA10291130 | PRJEB44396 |
| SAMEA10291122 | PRJEB44396 |
| SAMEA10291166 | PRJEB44396 |
| SAMEA10291118 | PRJEB44396 |
| SAMEA10291074 | PRJEB44396 |
| SAMEA10291073 | PRJEB44396 |
| SAMEA10291199 | PRJEB44396 |
| SAMEA10291174 | PRJEB44396 |
| SAMEA10291153 | PRJEB44396 |
| SAMEA10291102 | PRJEB44396 |

| | |
|---|---|
| SAMEA10291198 | PRJEB44396 |
| SAMEA10291094 | PRJEB44396 |
| SAMEA10291170 | PRJEB44396 |
| SAMEA10291088 | PRJEB44396 |
| SAMEA10291062 | PRJEB44396 |
| SAMEA10291081 | PRJEB44396 |
| SAMEA10291080 | PRJEB44396 |
| SAMEA10290991 | PRJEB44396 |
| SAMEA10290988 | PRJEB44396 |
| SAMEA10290984 | PRJEB44396 |
| SAMEA10291013 | PRJEB44396 |
| SAMEA10291010 | PRJEB44396 |
| SAMEA10291009 | PRJEB44396 |
| SAMEA10290976 | PRJEB44396 |
| SAMEA10290924 | PRJEB44396 |
| SAMEA10290975 | PRJEB44396 |
| SAMEA10291043 | PRJEB44396 |
| SAMEA10291007 | PRJEB44396 |
| SAMEA10290973 | PRJEB44396 |
| SAMEA10290896 | PRJEB44396 |
| SAMEA10290922 | PRJEB44396 |
| SAMEA10290970 | PRJEB44396 |
| SAMEA10291005 | PRJEB44396 |
| SAMEA10291047 | PRJEB44396 |
| SAMEA10291032 | PRJEB44396 |
| SAMEA10291042 | PRJEB44396 |
| SAMEA10290963 | PRJEB44396 |
| SAMEA10290960 | PRJEB44396 |
| SAMEA10290914 | PRJEB44396 |
| SAMEA10290955 | PRJEB44396 |
| SAMEA10290951 | PRJEB44396 |
| SAMEA10290998 | PRJEB44396 |
| SAMEA10290908 | PRJEB44396 |
| SAMEA10290948 | PRJEB44396 |
| SAMEA10290947 | PRJEB44396 |
| SAMEA10290995 | PRJEB44396 |
| SAMEA10291039 | PRJEB44396 |
| SAMEA10291026 | PRJEB44396 |
| SAMEA10291041 | PRJEB44396 |
| SAMEA10290994 | PRJEB44396 |
| SAMEA10291015 | PRJEB44396 |
| SAMEA10290993 | PRJEB44396 |
| SAMEA10290901 | PRJEB44396 |
| SAMEA10290992 | PRJEB44396 |
| SAMEA10290936 | PRJEB44396 |
| SAMEA10290898 | PRJEB44396 |
| SAMEA10290839 | PRJEB44396 |
| SAMEA10290872 | PRJEB44396 |

| | |
|---|---|
| SAMEA10290884 | PRJEB44396 |
| SAMEA10290870 | PRJEB44396 |
| SAMEA10290869 | PRJEB44396 |
| SAMEA10290879 | PRJEB44396 |
| SAMEA10290866 | PRJEB44396 |
| SAMEA10290864 | PRJEB44396 |
| SAMEA10290862 | PRJEB44396 |
| SAMEA10290877 | PRJEB44396 |
| SAMEA10290861 | PRJEB44396 |
| SAMEA10290860 | PRJEB44396 |
| SAMEA10290859 | PRJEB44396 |
| SAMEA10290858 | PRJEB44396 |
| SAMEA10290856 | PRJEB44396 |
| SAMEA10290831 | PRJEB44396 |
| SAMEA10290849 | PRJEB44396 |
| SAMEA10290848 | PRJEB44396 |
| SAMEA10290847 | PRJEB44396 |
| SAMEA10290887 | PRJEB44396 |
| SAMEA10290827 | PRJEB44396 |
| SAMEA10290888 | PRJEB44396 |
| SAMEA10290797 | PRJEB44396 |
| SAMEA10290803 | PRJEB44396 |
| SAMEA10290802 | PRJEB44396 |
| SAMEA10290826 | PRJEB44396 |
| SAMEA10290796 | PRJEB44396 |
| SAMEA10290825 | PRJEB44396 |
| SAMEA10290822 | PRJEB44396 |
| SAMEA10290817 | PRJEB44396 |
| SAMEA10290810 | PRJEB44396 |
| SAMEA10290807 | PRJEB44396 |
| SAMEA10290782 | PRJEB44396 |
| SAMEA10290544 | PRJEB44396 |
| SAMEA10290593 | PRJEB44396 |
| SAMEA10290610 | PRJEB44396 |
| SAMEA10290536 | PRJEB44396 |
| SAMEA10290535 | PRJEB44396 |
| SAMEA10290588 | PRJEB44396 |
| SAMEA10290532 | PRJEB44396 |
| SAMEA10290531 | PRJEB44396 |
| SAMEA10290583 | PRJEB44396 |
| SAMEA10290570 | PRJEB44396 |
| SAMEA10290568 | PRJEB44396 |
| SAMEA10290565 | PRJEB44396 |
| SAMEA10290501 | PRJEB44396 |
| SAMEA10253765 | PRJEB47798 |
| SAMEA10271074 | PRJEB47798 |
| SAMEA10271057 | PRJEB47798 |
| SAMEA10253708 | PRJEB47798 |

| | |
|---|---|
| SAMEA12009183 | PRJEB47798 |
| SAMEA10253774 | PRJEB47798 |
| SAMEA12009184 | PRJEB47798 |
| SAMEA10271085 | PRJEB47798 |
| SAMEA10253724 | PRJEB47798 |
| SAMEA12009185 | PRJEB47798 |
| SAMEA10253730 | PRJEB47798 |
| SAMEA10253788 | PRJEB47798 |
| SAMEA12009186 | PRJEB47798 |
| SAMEA12009187 | PRJEB47798 |
| SAMEA10253812 | PRJEB47798 |
| SAMEA10253764 | PRJEB47798 |
| SAMEA12009188 | PRJEB47798 |
| SAMEA10253750 | PRJEB47798 |
| SAMEA12009189 | PRJEB47798 |
| SAMEA10253715 | PRJEB47798 |
| SAMEA10253756 | PRJEB47798 |
| SAMEA10253864 | PRJEB47798 |
| SAMEA10253816 | PRJEB47798 |
| SAMEA10271075 | PRJEB47798 |
| SAMEA10253820 | PRJEB47798 |
| SAMEA10253768 | PRJEB47798 |
| SAMEA10253727 | PRJEB47798 |
| SAMEA10253736 | PRJEB47798 |
| SAMEA10253731 | PRJEB47798 |
| SAMEA10253748 | PRJEB47798 |
| SAMEA10253721 | PRJEB47798 |
| SAMEA10253728 | PRJEB47798 |
| SAMEA10253804 | PRJEB47798 |
| SAMEA10253819 | PRJEB47798 |
| SAMEA10253808 | PRJEB47798 |
| SAMEA10260231 | PRJEB47798 |
| SAMEA10260310 | PRJEB47798 |
| SAMEA10260295 | PRJEB47798 |
| SAMEA10260307 | PRJEB47798 |
| SAMEA10260239 | PRJEB47798 |
| SAMEA12009190 | PRJEB47798 |
| SAMEA10260316 | PRJEB47798 |
| SAMEA10260303 | PRJEB47798 |
| SAMEA12009191 | PRJEB47798 |
| SAMEA12009192 | PRJEB47798 |
| SAMEA12009193 | PRJEB47798 |
| SAMEA12009194 | PRJEB47798 |
| SAMEA12009195 | PRJEB47798 |
| SAMEA10271527 | PRJEB47798 |
| SAMEA10271530 | PRJEB47798 |
| SAMEA12009196 | PRJEB47798 |
| SAMEA10271511 | PRJEB47798 |

| | |
|---|---|
| SAMEA10271509 | PRJEB47798 |
| SAMEA10271526 | PRJEB47798 |
| SAMEA10271538 | PRJEB47798 |
| SAMEA10271507 | PRJEB47798 |
| SAMEA10271040 | PRJEB47798 |
| SAMEA12009197 | PRJEB47798 |
| SAMEA12009198 | PRJEB47798 |
| SAMEA10271043 | PRJEB47798 |
| SAMEA12009199 | PRJEB47798 |
| SAMEA10271550 | PRJEB47798 |
| SAMEA12009200 | PRJEB47798 |
| SAMEA10271553 | PRJEB47798 |
| SAMEA10271551 | PRJEB47798 |
| SAMEA10271049 | PRJEB47798 |
| SAMEA12009201 | PRJEB47798 |
| SAMEA10271052 | PRJEB47798 |
| SAMEA12009202 | PRJEB47798 |
| SAMEA10253840 | PRJEB47798 |
| SAMEA10253760 | PRJEB47798 |
| SAMEA10253725 | PRJEB47798 |
| SAMEA10253770 | PRJEB47798 |
| SAMEA10253755 | PRJEB47798 |
| SAMEA10253803 | PRJEB47798 |
| SAMEA10253779 | PRJEB47798 |
| SAMEA10253854 | PRJEB47798 |
| SAMEA10253801 | PRJEB47798 |
| SAMEA10253792 | PRJEB47798 |
| SAMEA10253836 | PRJEB47798 |
| SAMEA10253746 | PRJEB47798 |
| SAMEA10253862 | PRJEB47798 |
| SAMEA12009203 | PRJEB47798 |
| SAMEA10253845 | PRJEB47798 |
| SAMEA10253729 | PRJEB47798 |
| SAMEA10253815 | PRJEB47798 |
| SAMEA10253807 | PRJEB47798 |
| SAMEA12009204 | PRJEB47798 |
| SAMEA10253811 | PRJEB47798 |
| SAMEA10253800 | PRJEB47798 |
| SAMEA12009205 | PRJEB47798 |
| SAMEA10253777 | PRJEB47798 |
| SAMEA12009206 | PRJEB47798 |
| SAMEA10253844 | PRJEB47798 |
| SAMEA10253796 | PRJEB47798 |
| SAMEA10253810 | PRJEB47798 |
| SAMEA10253734 | PRJEB47798 |
| SAMEA10253817 | PRJEB47798 |
| SAMEA10253744 | PRJEB47798 |
| SAMEA10253758 | PRJEB47798 |

| | |
|---|---|
| SAMEA10253821 | PRJEB47798 |
| SAMEA10253710 | PRJEB47798 |
| SAMEA10253814 | PRJEB47798 |
| SAMEA10253754 | PRJEB47798 |
| SAMEA10253795 | PRJEB47798 |
| SAMEA12009207 | PRJEB47798 |
| SAMEA10253752 | PRJEB47798 |
| SAMEA10253809 | PRJEB47798 |
| SAMEA10270430 | PRJEB47798 |
| SAMEA10270449 | PRJEB47798 |
| SAMEA12009208 | PRJEB47798 |
| SAMEA10270541 | PRJEB47798 |
| SAMEA10270427 | PRJEB47798 |
| SAMEA10270412 | PRJEB47798 |
| SAMEA10270578 | PRJEB47798 |
| SAMEA10270556 | PRJEB47798 |
| SAMEA10270411 | PRJEB47798 |
| SAMEA10270505 | PRJEB47798 |
| SAMEA10270446 | PRJEB47798 |
| SAMEA10270555 | PRJEB47798 |
| SAMEA10270464 | PRJEB47798 |
| SAMEA10253719 | PRJEB47798 |
| SAMEA10270577 | PRJEB47798 |
| SAMEA10270445 | PRJEB47798 |
| SAMEA10270501 | PRJEB47798 |
| SAMEA12009209 | PRJEB47798 |
| SAMEA10270538 | PRJEB47798 |
| SAMEA10253848 | PRJEB47798 |
| SAMEA10253784 | PRJEB47798 |
| SAMEA10253847 | PRJEB47798 |
| SAMEA10253711 | PRJEB47798 |
| SAMEA10253789 | PRJEB47798 |
| SAMEA10253753 | PRJEB47798 |
| SAMEA10253794 | PRJEB47798 |
| SAMEA10253853 | PRJEB47798 |
| SAMEA10271175 | PRJEB47798 |
| SAMEA10271157 | PRJEB47798 |
| SAMEA10270500 | PRJEB47798 |
| SAMEA10270483 | PRJEB47798 |
| SAMEA10270484 | PRJEB47798 |
| SAMEA10270481 | PRJEB47798 |
| SAMEA10270480 | PRJEB47798 |
| SAMEA10270423 | PRJEB47798 |
| SAMEA12009210 | PRJEB47798 |
| SAMEA10270442 | PRJEB47798 |
| SAMEA12009211 | PRJEB47798 |
| SAMEA10270576 | PRJEB47798 |
| SAMEA10270441 | PRJEB47798 |

| | |
|---|---|
| SAMEA10270479 | PRJEB47798 |
| SAMEA12009212 | PRJEB47798 |
| SAMEA12009213 | PRJEB47798 |
| SAMEA10270574 | PRJEB47798 |
| SAMEA12009214 | PRJEB47798 |
| SAMEA10270572 | PRJEB47798 |
| SAMEA10270552 | PRJEB47798 |
| SAMEA10270458 | PRJEB47798 |
| SAMEA10270536 | PRJEB47798 |
| SAMEA10270438 | PRJEB47798 |
| SAMEA10270476 | PRJEB47798 |
| SAMEA10270518 | PRJEB47798 |
| SAMEA10270421 | PRJEB47798 |
| SAMEA10270517 | PRJEB47798 |
| SAMEA10271164 | PRJEB47798 |
| SAMEA10271145 | PRJEB47798 |
| SAMEA10271163 | PRJEB47798 |
| SAMEA10271078 | PRJEB47798 |
| SAMEA10271155 | PRJEB47798 |
| SAMEA12009215 | PRJEB47798 |
| SAMEA10271133 | PRJEB47798 |
| SAMEA10271103 | PRJEB47798 |
| SAMEA12009216 | PRJEB47798 |
| SAMEA10271165 | PRJEB47798 |
| SAMEA10271142 | PRJEB47798 |
| SAMEA10271129 | PRJEB47798 |
| SAMEA12009217 | PRJEB47798 |
| SAMEA10271153 | PRJEB47798 |
| SAMEA10271132 | PRJEB47798 |
| SAMEA10260299 | PRJEB47798 |
| SAMEA10260258 | PRJEB47798 |
| SAMEA12009218 | PRJEB47798 |
| SAMEA10260284 | PRJEB47798 |
| SAMEA10260264 | PRJEB47798 |
| SAMEA10260263 | PRJEB47798 |
| SAMEA10260267 | PRJEB47798 |
| SAMEA10271107 | PRJEB47798 |
| SAMEA10271160 | PRJEB47798 |
| SAMEA12009219 | PRJEB47798 |
| SAMEA10271121 | PRJEB47798 |
| SAMEA10271138 | PRJEB47798 |
| SAMEA10271545 | PRJEB47798 |
| SAMEA10271036 | PRJEB47798 |
| SAMEA12009220 | PRJEB47798 |
| SAMEA12009221 | PRJEB47798 |
| SAMEA10271547 | PRJEB47798 |
| SAMEA12009222 | PRJEB47798 |
| SAMEA10271544 | PRJEB47798 |

| | |
|---|---|
| SAMEA10260280 | PRJEB47798 |
| SAMEA10260247 | PRJEB47798 |
| SAMEA12009223 | PRJEB47798 |
| SAMEA10260240 | PRJEB47798 |
| SAMEA10260312 | PRJEB47798 |
| SAMEA10260287 | PRJEB47798 |
| SAMEA10260259 | PRJEB47798 |
| SAMEA10260245 | PRJEB47798 |
| SAMEA10260285 | PRJEB47798 |
| SAMEA10260260 | PRJEB47798 |
| SAMEA10260304 | PRJEB47798 |
| SAMEA12009224 | PRJEB47798 |
| SAMEA10260302 | PRJEB47798 |
| SAMEA10260308 | PRJEB47798 |
| SAMEA10260265 | PRJEB47798 |
| SAMEA12009225 | PRJEB47798 |
| SAMEA12009226 | PRJEB47798 |
| SAMEA10270453 | PRJEB47798 |
| SAMEA10270475 | PRJEB47798 |
| SAMEA12009227 | PRJEB47798 |
| SAMEA10270437 | PRJEB47798 |
| SAMEA10271180 | PRJEB47798 |
| SAMEA10253782 | PRJEB47798 |
| SAMEA10253793 | PRJEB47798 |
| SAMEA10253716 | PRJEB47798 |
| SAMEA10253757 | PRJEB47798 |
| SAMEA10271178 | PRJEB47798 |
| SAMEA10271120 | PRJEB47798 |
| SAMEA10271170 | PRJEB47798 |
| SAMEA10271156 | PRJEB47798 |
| SAMEA10271122 | PRJEB47798 |
| SAMEA10271126 | PRJEB47798 |
| SAMEA10271119 | PRJEB47798 |
| SAMEA10271151 | PRJEB47798 |
| SAMEA10271117 | PRJEB47798 |
| SAMEA10271137 | PRJEB47798 |
| SAMEA10271060 | PRJEB47798 |
| SAMEA10271110 | PRJEB47798 |
| SAMEA10271169 | PRJEB47798 |
| SAMEA10253850 | PRJEB47798 |
| SAMEA10271177 | PRJEB47798 |
| SAMEA10253837 | PRJEB47798 |
| SAMEA10271099 | PRJEB47798 |
| SAMEA10271091 | PRJEB47798 |
| SAMEA10271070 | PRJEB47798 |
| SAMEA12009228 | PRJEB47798 |
| SAMEA10270569 | PRJEB47798 |
| SAMEA10253858 | PRJEB47798 |

| | |
|---|---|
| SAMEA10270531 | PRJEB47798 |
| SAMEA10270493 | PRJEB47798 |
| SAMEA10260296 | PRJEB47798 |
| SAMEA10260237 | PRJEB47798 |
| SAMEA10260244 | PRJEB47798 |
| SAMEA12009229 | PRJEB47798 |
| SAMEA10260274 | PRJEB47798 |
| SAMEA10271130 | PRJEB47798 |
| SAMEA10271055 | PRJEB47798 |
| SAMEA10271058 | PRJEB47798 |
| SAMEA10271109 | PRJEB47798 |
| SAMEA10271125 | PRJEB47798 |
| SAMEA10271124 | PRJEB47798 |
| SAMEA10271108 | PRJEB47798 |
| SAMEA10271059 | PRJEB47798 |
| SAMEA10271105 | PRJEB47798 |
| SAMEA10271081 | PRJEB47798 |
| SAMEA10271063 | PRJEB47798 |
| SAMEA10271073 | PRJEB47798 |
| SAMEA10271100 | PRJEB47798 |
| SAMEA10271094 | PRJEB47798 |
| SAMEA12009230 | PRJEB47798 |
| SAMEA10271123 | PRJEB47798 |
| SAMEA10271090 | PRJEB47798 |
| SAMEA12009231 | PRJEB47798 |
| SAMEA10271096 | PRJEB47798 |
| SAMEA10271072 | PRJEB47798 |
| SAMEA10271089 | PRJEB47798 |
| SAMEA10271106 | PRJEB47798 |
| SAMEA10271064 | PRJEB47798 |
| SAMEA12009232 | PRJEB47798 |
| SAMEA10271093 | PRJEB47798 |
| SAMEA10270407 | PRJEB47798 |
| SAMEA10270416 | PRJEB47798 |
| SAMEA10270546 | PRJEB47798 |
| SAMEA10270514 | PRJEB47798 |
| SAMEA10270406 | PRJEB47798 |
| SAMEA10270452 | PRJEB47798 |
| SAMEA10270405 | PRJEB47798 |
| SAMEA10270513 | PRJEB47798 |
| SAMEA10270565 | PRJEB47798 |
| SAMEA10270512 | PRJEB47798 |
| SAMEA10270473 | PRJEB47798 |
| SAMEA8922088 | PRJEB43166 |
| SAMEA8922114 | PRJEB43166 |
| SAMEA8922109 | PRJEB43166 |
| SAMEA8922102 | PRJEB43166 |
| SAMEA8922091 | PRJEB43166 |

| | |
|---|---|
| SAMEA8922056 | PRJEB43166 |
| SAMEA8922046 | PRJEB43166 |
| SAMEA8922104 | PRJEB43166 |
| SAMEA8922092 | PRJEB43166 |
| SAMEA8922083 | PRJEB43166 |
| SAMEA8922078 | PRJEB43166 |
| SAMEA8922076 | PRJEB43166 |
| SAMEA8922037 | PRJEB43166 |
| SAMEA8922036 | PRJEB43166 |
| SAMEA8922094 | PRJEB43166 |
| SAMEA8922075 | PRJEB43166 |
| SAMEA8922098 | PRJEB43166 |
| SAMEA8922117 | PRJEB43166 |
| SAMEA8922113 | PRJEB43166 |
| SAMEA8922112 | PRJEB43166 |
| SAMEA8922110 | PRJEB43166 |
| SAMEA8922108 | PRJEB43166 |
| SAMEA8922097 | PRJEB43166 |
| SAMEA8922096 | PRJEB43166 |
| SAMEA8922095 | PRJEB43166 |
| SAMEA8922093 | PRJEB43166 |
| SAMEA8922089 | PRJEB43166 |
| SAMEA8922087 | PRJEB43166 |
| SAMEA8922086 | PRJEB43166 |
| SAMEA8922085 | PRJEB43166 |
| SAMEA8922084 | PRJEB43166 |
| SAMEA8922082 | PRJEB43166 |
| SAMEA8922080 | PRJEB43166 |
| SAMEA8922079 | PRJEB43166 |
| SAMEA8922074 | PRJEB43166 |
| SAMEA8922073 | PRJEB43166 |
| SAMEA8922041 | PRJEB43166 |
| SAMEA8922090 | PRJEB43166 |
| SAMEA8922081 | PRJEB43166 |
| SAMEA8921680 | PRJEB43166 |
| SAMEA8921678 | PRJEB43166 |
| SAMEA8921676 | PRJEB43166 |
| SAMEA8921284 | PRJEB43166 |
| SAMEA8921282 | PRJEB43166 |
| SAMEA8921280 | PRJEB43166 |
| SAMEA8921277 | PRJEB43166 |
| SAMEA8921271 | PRJEB43166 |
| SAMEA8921266 | PRJEB43166 |
| SAMEA8921263 | PRJEB43166 |
| SAMEA8921260 | PRJEB43166 |
| SAMEA8921254 | PRJEB43166 |
| SAMEA8921247 | PRJEB43166 |
| SAMEA8921245 | PRJEB43166 |

| | |
|---|---|
| SAMEA8921232 | PRJEB43166 |
| SAMEA8921229 | PRJEB43166 |
| SAMEA8921228 | PRJEB43166 |
| SAMEA8921221 | PRJEB43166 |
| SAMEA8921216 | PRJEB43166 |
| SAMEA8921215 | PRJEB43166 |
| SAMEA8921214 | PRJEB43166 |
| SAMEA8921211 | PRJEB43166 |
| SAMEA8921206 | PRJEB43166 |
| SAMEA8921204 | PRJEB43166 |
| SAMEA8921608 | PRJEB43166 |
| SAMEA8921378 | PRJEB43166 |
| SAMEA8921366 | PRJEB43166 |
| SAMEA8921203 | PRJEB43166 |
| SAMEA8921201 | PRJEB43166 |
| SAMEA8921200 | PRJEB43166 |
| SAMEA8921199 | PRJEB43166 |
| SAMEA8921198 | PRJEB43166 |
| SAMEA8921197 | PRJEB43166 |
| SAMEA8921192 | PRJEB43166 |
| SAMEA8921187 | PRJEB43166 |
| SAMEA8921186 | PRJEB43166 |
| SAMEA8921185 | PRJEB43166 |
| SAMEA8921181 | PRJEB43166 |
| SAMEA8921179 | PRJEB43166 |
| SAMEA8921165 | PRJEB43166 |
| SAMEA8921163 | PRJEB43166 |
| SAMEA8921162 | PRJEB43166 |
| SAMEA8921160 | PRJEB43166 |
| SAMEA8921158 | PRJEB43166 |
| SAMEA8921156 | PRJEB43166 |
| SAMEA8921155 | PRJEB43166 |
| SAMEA8921154 | PRJEB43166 |
| SAMEA8921153 | PRJEB43166 |
| SAMEA8921152 | PRJEB43166 |
| SAMEA8921151 | PRJEB43166 |
| SAMEA8921150 | PRJEB43166 |
| SAMEA8921138 | PRJEB43166 |
| SAMEA8921135 | PRJEB43166 |
| SAMEA8921132 | PRJEB43166 |
| SAMEA8921130 | PRJEB43166 |
| SAMEA8922043 | PRJEB43166 |
| SAMEA8922115 | PRJEB43166 |
| SAMEA8922107 | PRJEB43166 |
| SAMEA8922067 | PRJEB43166 |
| SAMEA8922100 | PRJEB43166 |
| SAMEA8922099 | PRJEB43166 |
| SAMEA8922105 | PRJEB43166 |

| | |
|---|---|
| SAMEA8922070 | PRJEB43166 |
| SAMEA8921372 | PRJEB43166 |
