## Supplementary Table S2 for "Assessing the impact of SARS-CoV-2 lineages and mutations on patient survival"

Nucleotide mutations eligible for causal analysis. The first column is the mutation name; the second is the position; the third column, labeled as CDS, is the protein affected; the fourth column is the amino acid mutation name; the fifth column is the number of variants bearing this mutation; and the following columns provide the values of the two approaches for hazard ratio estimation, the closed form, with the hazard ratio coefficient, SD, confidence intervals 5 and 95, the p-value and the FDR adjusted p-value, and the bootstrap approach with the HR coefficients (Boot. Statistic), bias, SD, confidence intervals 5 and 95 and the last column, labeled as Boot, indicates if significance is confirmed by bootstrap (T: true and F: false)

| Mutation name | position | CDS | AAc mutation name | cases | Theor coeff. | Theor. StD | Theor. CI05 | Theor. CI95 | Theor. P-val | Theor. P-val FDR | Boot. statistic | Boot bias | Boot. Std Err | Boot CI 05 | Boot CI 95 | Boot |
|---|---|---|---|---|---|---|---|---|---|---|---|---|---|---|---|---|
| 28274 | 28274 | N | N:M1M | 291 | 0.5614 | 0.1839 | 0.2010 | 0.9219 | 0.0023 | 0.0423 | 0.5614 | 0.0030 | 0.1940 | 0.1832 | 0.9322 | T |
| C5388A | 5388 | ORF1ab | ORF1ab:A1708D | 288 | 0.5213 | 0.1835 | 0.1616 | 0.8811 | 0.0045 | 0.0423 | 0.5213 | -0.0061 | 0.1873 | 0.1795 | 0.9136 | T |
| C23709T | 23709 | S | S:T716I | 294 | 0.5179 | 0.1827 | 0.1598 | 0.8761 | 0.0046 | 0.0423 | 0.5179 | -0.0058 | 0.1912 | 0.1450 | 0.8933 | T |
| T6954C | 6954 | ORF1ab | ORF1ab:I2230T | 284 | 0.5208 | 0.1843 | 0.1595 | 0.8821 | 0.0047 | 0.0423 | 0.5208 | 0.0037 | 0.1892 | 0.1345 | 0.8931 | T |
| C14676T | 14676 | ORF1ab | ORF1ab:P4803P | 288 | 0.5136 | 0.1834 | 0.1542 | 0.8731 | 0.0051 | 0.0423 | 0.5136 | 0.0019 | 0.1867 | 0.0840 | 0.8374 | T |
| C15279T | 15279 | ORF1ab | ORF1ab:H5004H | 288 | 0.5049 | 0.1824 | 0.1474 | 0.8625 | 0.0056 | 0.0423 | 0.5049 | -0.0107 | 0.1932 | 0.1358 | 0.8936 | T |
| C23604A | 23604 | S | S:P681H | 296 | 0.5033 | 0.1828 | 0.1450 | 0.8615 | 0.0059 | 0.0423 | 0.5033 | -0.0062 | 0.1902 | 0.1215 | 0.8759 | T |
| 11288,11297 | 11288 | ORF1ab | ORF1ab:del3675-3677 | 291 | 0.4957 | 0.1831 | 0.1368 | 0.8545 | 0.0068 | 0.0423 | 0.4957 | 0.0022 | 0.1906 | 0.1271 | 0.8695 | T |
| C23271A | 23271 | S | S:A570D | 289 | 0.4853 | 0.1821 | 0.1283 | 0.8422 | 0.0077 | 0.0423 | 0.4853 | -0.0093 | 0.1863 | 0.1368 | 0.8545 | T |
| G28280C | 28280 | N | N:D3L | 288 | 0.4856 | 0.1838 | 0.1254 | 0.8458 | 0.0082 | 0.0423 | 0.4856 | -0.0021 | 0.1858 | 0.1307 | 0.8725 | T |
| A28281T | 28281 | N | N:D3L | 288 | 0.4856 | 0.1838 | 0.1254 | 0.8458 | 0.0082 | 0.0423 | 0.4856 | -0.0021 | 0.1858 | 0.1307 | 0.8725 | T |
| T28282A | 28282 | N | N:D3L | 288 | 0.4856 | 0.1838 | 0.1254 | 0.8458 | 0.0082 | 0.0423 | 0.4856 | -0.0021 | 0.1858 | 0.1307 | 0.8725 | T |
| A28111G | 28111 | ORF8 | ORF8:Y73C | 288 | 0.4863 | 0.1846 | 0.1244 | 0.8481 | 0.0084 | 0.0423 | 0.4863 | -0.0050 | 0.1923 | 0.1458 | 0.9159 | T |
| C913T | 913 | ORF1ab | ORF1ab:S216S | 288 | 0.4805 | 0.1828 | 0.1221 | 0.8388 | 0.0086 | 0.0423 | 0.4805 | -0.0078 | 0.1878 | 0.1423 | 0.8535 | T |
| T24506G | 24506 | S | S:S982A | 290 | 0.4721 | 0.1829 | 0.1136 | 0.8306 | 0.0099 | 0.0442 | 0.4721 | -0.0002 | 0.1874 | 0.0670 | 0.8295 | T |
| C5986T | 5986 | ORF1ab | ORF1ab:F1907F | 288 | 0.4677 | 0.1822 | 0.1106 | 0.8247 | 0.0102 | 0.0442 | 0.4677 | -0.0043 | 0.1892 | 0.0976 | 0.8338 | T |
| C3267T | 3267 | ORF1ab | ORF1ab:T1001I | 290 | 0.4612 | 0.1818 | 0.1049 | 0.8175 | 0.0112 | 0.0454 | 0.4612 | 0.0015 | 0.1903 | 0.0775 | 0.8307 | T |
| G24914C | 24914 | S | S:D1118H | 288 | 0.4604 | 0.1832 | 0.1013 | 0.8194 | 0.0120 | 0.0459 | 0.4604 | 0.0073 | 0.1904 | 0.0846 | 0.8228 | T |
| 21766,21772 | 21766 | S | S:del69-70 | 280 | 0.4567 | 0.1836 | 0.0969 | 0.8164 | 0.0128 | 0.0467 | 0.4567 | 0.0027 | 0.1942 | 0.0495 | 0.8196 | T |
| G28048T | 28048 | ORF8 | ORF8:R52I | 289 | 0.4571 | 0.1852 | 0.0940 | 0.8201 | 0.0136 | 0.0468 | 0.4571 | -0.0038 | 0.1857 | 0.1008 | 0.8255 | T |
| C27972T | 27972 | ORF8 | ORF8:Q27* | 290 | 0.4515 | 0.1853 | 0.0883 | 0.8147 | 0.0148 | 0.0468 | 0.4515 | 0.0009 | 0.1875 | 0.0635 | 0.8169 | T |
| C28977T | 28977 | N | N:S235F | 290 | 0.4448 | 0.1827 | 0.0867 | 0.8029 | 0.0149 | 0.0468 | 0.4448 | -0.0038 | 0.1889 | 0.0531 | 0.8014 | T |

| | | | | | | | | | | | | | | | | |
|---|---|---|---|---|---|---|---|---|---|---|---|---|---|---|---|---|
| T16176C | 16176 | ORF1ab | ORF1ab:T5303T | 285 | 0.4390 | 0.1831 | 0.0801 | 0.7980 | 0.0165 | 0.0496 | 0.4390 | -0.0070 | 0.1855 | 0.0609 | 0.8025 | T |
| 21994,21997 | 21994 | S | S:Y144- | 270 | 0.4431 | 0.1874 | 0.0758 | 0.8104 | 0.0181 | 0.0519 | 0.4431 | 0.0050 | 0.1944 | 0.0663 | 0.8194 | T |
| A23063T | 23063 | S | S:N501Y | 273 | 0.4380 | 0.1901 | 0.0654 | 0.8107 | 0.0212 | 0.0586 | 0.4380 | -0.0080 | 0.1997 | 0.0314 | 0.8267 | T |
| C10833T | 10833 | ORF1ab | ORF1ab:A3523V | 26 | -0.9421 | 0.5182 | -1.9578 | 0.0736 | 0.0691 | 0.1833 | -0.9421 | -0.9488 | 4.1888 | -21.3190 | -0.0541 | T |
| C6968T | 6968 | ORF1ab | ORF1ab:L2235L | 71 | 0.4784 | 0.2707 | -0.0521 | 1.0089 | 0.0772 | 0.1972 | 0.4784 | -0.0019 | 0.2972 | -0.1305 | 1.0217 | F |
| G28882A | 28882 | NA | N:R203L | 315 | 0.3026 | 0.1807 | -0.0515 | 0.6567 | 0.0940 | 0.2235 | 0.3026 | 0.0027 | 0.1783 | -0.0528 | 0.6544 | F |
| G28883C | 28883 | N | N:G204R | 315 | 0.3026 | 0.1807 | -0.0515 | 0.6567 | 0.0940 | 0.2235 | 0.3026 | 0.0027 | 0.1783 | -0.0528 | 0.6544 | F |
| C22227T | 22227 | S | S:A222V | 73 | -0.4460 | 0.2846 | -1.0039 | 0.1118 | 0.1171 | 0.2606 | -0.4460 | -0.0298 | 0.3076 | -1.0691 | 0.1359 | F |
| G28881A | 28881 | N | N:R203L | 317 | 0.2829 | 0.1805 | -0.0709 | 0.6366 | 0.1171 | 0.2606 | 0.2829 | 0.0027 | 0.1777 | -0.0683 | 0.6288 | F |
| G21255C | 21255 | ORF1ab | ORF1ab:A6996A | 71 | -0.4022 | 0.2614 | -0.9145 | 0.1100 | 0.1238 | 0.2636 | -0.4022 | -0.0248 | 0.2838 | -1.0252 | 0.1064 | F |
| C222T | 222 | No ORF | No ORF | 32 | -0.7169 | 0.4687 | -1.6355 | 0.2016 | 0.1261 | 0.2636 | -0.7169 | -0.3770 | 2.4235 | -2.2067 | 0.1464 | F |
| G4300T | 4300 | ORF1ab | ORF1ab:V1345V | 5 | 1.3628 | 1.0428 | -0.6811 | 3.4067 | 0.1913 | 0.3882 | 1.3628 | -6.4484 | 12.3659 | -21.2936 | 21.1406 | F |
| C6286T | 6286 | ORF1ab | ORF1ab:T2007T | 74 | -0.3261 | 0.2598 | -0.8353 | 0.1831 | 0.2095 | 0.4058 | -0.3261 | -0.0211 | 0.2668 | -0.8521 | 0.1708 | F |
| G1820A | 1820 | ORF1ab | ORF1ab:G519S | 14 | 0.4874 | 0.3944 | -0.2856 | 1.2603 | 0.2165 | 0.4058 | 0.4874 | -0.0498 | 1.0898 | -0.5423 | 1.3784 | F |
| A17615G | 17615 | ORF1ab | ORF1ab:K5783R | 66 | 0.4193 | 0.3401 | -0.2473 | 1.0860 | 0.2176 | 0.4058 | 0.4193 | -0.0381 | 0.3852 | -0.3624 | 1.1394 | F |
| C2710T | 2710 | ORF1ab | ORF1ab:L815L | 12 | 0.6595 | 0.6019 | -0.5201 | 1.8392 | 0.2732 | 0.4948 | 0.6595 | -2.9978 | 7.6816 | -21.3015 | 2.1132 | F |
| G29734C | 29734 | No ORF | No ORF | 46 | 0.3360 | 0.3129 | -0.2772 | 0.9493 | 0.2828 | 0.4948 | 0.3360 | -0.0098 | 0.3497 | -0.3691 | 0.9999 | F |
| A23403G | 23403 | S | D614G | 616 | 0.2413 | 0.2266 | -0.2028 | 0.6854 | 0.2868 | 0.4948 | 0.2413 | 0.0166 | 0.2407 | -0.1949 | 0.7073 | F |
| T445C | 445 | ORF1ab | ORF1ab:V60V | 77 | -0.2506 | 0.2437 | -0.7283 | 0.2271 | 0.3038 | 0.5113 | -0.2506 | -0.0131 | 0.2589 | -0.7613 | 0.2766 | F |
| C28932T | 28932 | N, ORF9c | N:A220V, ORF9c:L67F | 71 | -0.2501 | 0.2557 | -0.7513 | 0.2511 | 0.3281 | 0.5119 | -0.2501 | -0.0230 | 0.2657 | -0.7788 | 0.2541 | F |
| C26801G | 26801 | M | M:L93L | 72 | -0.2469 | 0.2560 | -0.7487 | 0.2549 | 0.3349 | 0.5119 | -0.2469 | -0.0206 | 0.2745 | -0.8157 | 0.2896 | F |
| C14408T | 14408 | ORF1ab | ORF1ab:P4713L | 615 | 0.2193 | 0.2275 | -0.2266 | 0.6653 | 0.3350 | 0.5119 | 0.2193 | 0.0260 | 0.2498 | -0.2387 | 0.7457 | F |
| T29589C | 29589 | ORF10 | ORF10:F11S | 75 | 0.2732 | 0.2837 | -0.2830 | 0.8293 | 0.3357 | 0.5119 | 0.2732 | -0.0163 | 0.3053 | -0.4343 | 0.8070 | F |
| G29645T | 29645 | ORF10 | ORF10:V30L | 71 | -0.2428 | 0.2551 | -0.7428 | 0.2572 | 0.3413 | 0.5119 | -0.2428 | -0.0287 | 0.2715 | -0.7374 | 0.3042 | F |
| C25463A | 25463 | ORF3a, ORF3c | ORF3a:T24N, ORF3c:L3I | 12 | 0.5123 | 0.6054 | -0.6742 | 1.6989 | 0.3974 | 0.5779 | 0.5123 | -2.5029 | 7.1754 | -21.4067 | 2.4443 | F |
| C25714A | 25714 | ORF3a | ORF3a:L108I | 7 | -0.3967 | 0.4734 | -1.3246 | 0.5312 | 0.4021 | 0.5779 | -0.3967 | -2.5603 | 7.1736 | -21.9799 | 1.1567 | F |
| C16887T | 13887 | ORF1ab | ORF1ab:Y5540Y | 79 | 0.2138 | 0.2701 | -0.3156 | 0.7432 | 0.4286 | 0.5985 | 0.2138 | -0.0139 | 0.2854 | -0.3846 | 0.7693 | F |
| G12892A | 12892 | ORF1ab | ORF1ab:L4209L | 14 | -0.7716 | 0.9856 | -2.7034 | 1.1602 | 0.4337 | 0.5985 | -0.7716 | -7.0538 | 10.0145 | -21.5310 | 0.9286 | F |
| C8782T | 8782 | ORF1ab | ORF1ab:S2839S | 142 | -0.1813 | 0.2362 | -0.6443 | 0.2817 | 0.4428 | 0.5991 | -0.1813 | -0.0228 | 0.2408 | -0.6376 | 0.3152 | F |
| T28144C | 28144 | ORF8 | ORF8:L84S | 145 | -0.1690 | 0.2308 | -0.6214 | 0.2835 | 0.4642 | 0.6159 | -0.1690 | -0.0129 | 0.2394 | -0.6670 | 0.2782 | F |
| C241T | 241 | No ORF | No ORF | 624 | 0.1596 | 0.2238 | -0.2789 | 0.5982 | 0.4756 | 0.6192 | 0.1596 | 0.0151 | 0.2354 | -0.3052 | 0.6408 | F |

| | | | | | | | | | | | | | | | | |
|---|---|---|---|---|---|---|---|---|---|---|---|---|---|---|---|---|
| C3037T | 3037 | ORF1ab | ORF1ab:F924F | 624 | 0.1554 | 0.2237 | -0.2831 | 0.5939 | 0.4872 | 0.6225 | 0.1554 | 0.0186 | 0.2355 | -0.2878 | 0.6425 | F |
| C13671T | 13671 | ORF1ab | ORF1ab:F4468F | 7 | 0.2569 | 0.4288 | -0.5836 | 1.0974 | 0.5491 | 0.6889 | 0.2569 | -0.9971 | 4.8332 | -21.4026 | 1.7911 | F |
| C27944T | 27944 | ORF8 | ORF8:H17H | 26 | 0.1639 | 0.3605 | -0.5426 | 0.8705 | 0.6492 | 0.7999 | 0.1639 | -0.0351 | 0.4125 | -0.7527 | 0.8791 | F |
| C28863T | 28863 | N, ORF9c | N:S197L, ORF9c:Q44- | 122 | -0.0885 | 0.2420 | -0.5628 | 0.3857 | 0.7145 | 0.8649 | -0.0885 | -0.0135 | 0.2586 | -0.6829 | 0.3750 | F |
| C28657T | 28657 | N | N:D128D | 120 | -0.0692 | 0.2399 | -0.5394 | 0.4010 | 0.7730 | 0.8987 | -0.0692 | -0.0270 | 0.2406 | -0.5288 | 0.4204 | F |
| C5833G | 5833 | ORF1ab | ORF1ab:S1856S | 6 | -0.1089 | 0.4240 | -0.9399 | 0.7221 | 0.7973 | 0.8987 | -0.1089 | -2.3846 | 6.9040 | -21.5059 | 1.8958 | F |
| T6552C | 6552 | ORF1ab | ORF1ab:M2096T | 12 | 0.2084 | 0.8795 | -1.5154 | 1.9322 | 0.8127 | 0.8987 | 0.2084 | -7.5357 | 10.3159 | -21.1259 | 1.7815 | F |
| G25979T | 25979 | ORF3a | ORF3a:G196V | 121 | -0.0537 | 0.2408 | -0.5258 | 0.4183 | 0.8235 | 0.8987 | -0.0537 | -0.0180 | 0.2527 | -0.5356 | 0.4367 | F |
| 21766,21771 | 21766 | S | S:del69 | 7 | 0.1781 | 0.8144 | -1.4181 | 1.7744 | 0.8269 | 0.8987 | 0.1781 | -7.4589 | 10.5512 | -21.3706 | 3.0984 | F |
| A20268G | 20268 | ORF1ab | ORF1ab:L6667L | 165 | 0.0417 | 0.2044 | -0.3589 | 0.4422 | 0.8383 | 0.8987 | 0.0417 | -0.0081 | 0.2132 | -0.3802 | 0.4344 | F |
| C14805T | 14805 | ORF1ab | ORF1ab:Y4846Y | 121 | -0.0440 | 0.2401 | -0.5146 | 0.4266 | 0.8546 | 0.8987 | -0.0440 | -0.0082 | 0.2502 | -0.6118 | 0.4066 | F |
| G25647T | 25647 | ORF3a, ORF3d | ORF3a_L85F, ORF3d:V42F | 12 | 0.1429 | 0.8582 | -1.5391 | 1.8249 | 0.8678 | 0.8987 | 0.1429 | -7.5487 | 10.3007 | -21.1876 | 1.8360 | F |
| G26262A | 26262 | M | M:S6S | 12 | 0.1431 | 0.8606 | -1.5436 | 1.8299 | 0.8679 | 0.8987 | 0.1431 | -7.0384 | 10.1695 | -21.2036 | 1.5148 | F |
| C28725T | 28725 | N | N:P151L | 12 | 0.1381 | 0.8618 | -1.5509 | 1.8272 | 0.8726 | 0.8987 | 0.1381 | -7.4038 | 10.2820 | -21.1775 | 1.8618 | F |
| G21974T | 21974 | S | S:D138Y | 12 | -0.0374 | 0.3523 | -0.7280 | 0.6531 | 0.9154 | 0.9288 | -0.0374 | -1.4007 | 5.2867 | -21.3816 | 0.8658 | F |
| T9477A | 9477 | ORF1ab | ORF1ab:F3071Y | 116 | -0.0138 | 0.2397 | -0.4836 | 0.4560 | 0.9540 | 0.9540 | -0.0138 | -0.0046 | 0.2531 | -0.5382 | 0.4574 | F |
