## Supplementary material for "Assessing the impact of SARS-CoV-2 lineages and mutations on patient survival": The Andalusian COVID-19 sequencing initiative

**Monica Perez-Alegre, Eloisa Andújar**: Centro Andaluz de Biología Molecular y Medicina Regenerativa CABIMER, Universidad de Sevilla-CSIC-Universidad Pablo de Olavide, Sevilla, Spain.

**Matilde Palanca Gimenez**: Hospital Poniente de Almería, El Ejido, Almería, Spain

**Manuel Rodríguez Maresca**: Hospital Torrecárdenas, Almería, Spain

**Manuel A. Rodríguez Iglesias**: Hospital Puerta del Mar, Cádiz, Spain

**Manuel Causse del Río, Cristina Riazzo, Luis Martínez-Martínez**: Hospital Universitario Reina Sofía, Córdoba, Spain

**Francisco Franco Álvarez De Luna**: Hospital Juan Ramón Jiménez, Huelva, Spain

**Carolina Roldán Fontana**: Complejo Hospitalario de Jaén, Jaén, Spain

**María Dolores López Prieto**: Hospital de Jerez, Cádiz, Spain

**Maria Luisa Hortas, Fernando Fernandez Sanchez**: Hospital Costa del Sol, Málaga, Spain

**Begoña Palop Borrás**: Hospital Regional, Málaga, Spain

**Isabel Viciana**: Hospital Virgen de la Victoria, Málaga, Spain

**Alvaro Pascual**: Hospital Virgen de la Macarena, Sevilla; Institute of Biomedicine of Seville (IBIS), Hospital Virgen del Rocio. 41013. Sevilla. Spain

**Ángel Rodríguez Villodres**: Hospital Universitario Virgen del Rocío, Sevilla, Spain

**Samuel Bernal Martinez, Estrella Martin Mazuelos**: Unidad Clinica de Enfermedades Infecciosas y Microbiología (UCEIM), H.U. Virgen de Valme, Sevilla

**Inés Ruiz Molina**: Hospital Punta Europa, Cádiz, Spain

**Natalia Chueca, Ana Fuentes**: Servicio de Microbiología. Unidad Clínica Enfermedades Infecciosas, Microbiología y Medicina Preventiva. Hospital Universitario Virgen del Rocio. Sevilla. Spain
